# Strengthening Post-Market Vaccine Safety Surveillance Globally: An Interpretive Description Study from Kenya, South Africa, and Canada

**DOI:** 10.64898/2026.03.11.26347757

**Authors:** Anila Naz AliSher, Shabnam Shaik, Nellie Myburgh, Nomsa Ndaba, Alex Hinga, Winnie Okore, Samuel Sang, Salima Siraj, Thabisile Qwabi, Kimberley Gutu, Allan Matano, Antonia Di Castri, Sonali Kochhar, Eunice Wangeci Kagucia, Ziyaad Dangor, Clare L. Cutland, Karina A. Top, the International Network of Special Immunization Services (INSIS) Investigators

## Abstract

**Background:** Post-market vaccine safety surveillance and research are essential to detect and evaluate rare adverse events following immunization (AEFIs) not identified in pre-licensure clinical trials, such as myocarditis post-COVID-19 vaccination. AEFI surveillance infrastructure varies between jurisdictions. Factors that enable or hinder AEFI investigation across diverse settings are not well understood.

**Objective:** To examine clinical, social, and structural/system factors that enable AEFI case identification, reporting, data and biosample collection in high-income countries (HICs) and low-and middle-income countries (LMICs).

**Methods:** We conducted a qualitative study informed by Interpretive Description in Kenya, South Africa, and Canada. Participants were recruited using purposive and snowball sampling from three groups: (1) key informants with leadership roles in AEFI surveillance and research, (2) healthcare workers, research and laboratory staff involved in AEFI investigation and research, and (3) past participants in vaccine studies. Data were collected through semi-structured interviews and focus group discussions. Thematic analysis was conducted.

**Results:** Eighteen key informants, 47 healthcare workers and research staff, and 27 past research participants were enrolled. Facilitators and barriers were identified across two domains: AEFI surveillance and investigation, and vaccine safety research participation. AEFI surveillance and investigation were shaped by trust in system responsiveness, beliefs and awareness around AEFI reporting, digital innovation, and implementation gaps. Research participation was shaped by altruism and social influence, logistics and research infrastructure, and institutional policies and privacy. Facilitators to AEFI surveillance and research included established policies and procedures for AEFI surveillance, digital tools, emphatic communication, and convenient research processes. Barriers included limitations in workforce capacity, diagnostic testing and funding (especially in LMICs), and cumbersome research approval processes.

**Conclusions:** Findings underscore the need for both globally harmonized standards and locally tailored strategies to support vaccine safety surveillance and research in LMICs and HICs that are patient/participant-centered and include sustainable investments in infrastructure and workforce capacity.

## Introduction

Effective vaccines against COVID-19 were developed with unprecedented speed and efficiency. As with some previous mass vaccine campaigns [1], rare adverse events following immunization (AEFIs) were identified after COVID-19 vaccines were introduced, including thrombosis with thrombocytopenia syndrome (TTS) associated with the adenoviral vector COVID-19 vaccines (e.g, ChAdOx1-S, AstraZeneca [Vaxrevia], Ad26.COV-2S, Janssen) and myocarditis and pericarditis associated with mRNA COVID-19 vaccines (i.e., BNT162b2, Pfizer-BioNTech [Comirnaty], mRNA-1273, Moderna [Spikevax]) [2–10].

The rarity of these AEFIs (occurring in fewer than 1-10 per 100,000 vaccinees) makes them challenging to investigate, as cases present sporadically in different places, times, and after different vaccines. Moreover, the incidence of an AEFI and the background rate of the adverse event in the population may differ by vaccine product, age group, sex, ancestry, geography and other factors, which can complicate detection of safety signals [7–11]. The capacity to diagnose and manage patients with AEFIs, as well as accessibility of vaccine and clinical information for surveillance purposes, also differ between health systems and jurisdictions, leading to differences in AEFI reporting rates and health outcomes [7,8,11]. It is therefore essential to have surveillance systems in place to detect and investigate AEFIs in low- and middle-income countries (LMICs) as well as in high-income countries (HICs), to fully characterize the safety profile of a vaccine across different settings and population groups.

However, to truly advance vaccine safety science and reduce the potential for AEFIs, we need to investigate the biological mechanisms and risk factors underpinning AEFIs such as COVID-19 vaccine-associated TTS and myocarditis [12]. This requires the timely, coordinated collection of large numbers of biosamples from post-vaccination cases and vaccinated controls worldwide, using harmonized protocols that enable analysis on common platforms [13]. To date, no single country has been able to collect enough biosamples on its own to conduct highly powered analyses into biomarker risk factors for AEFIs [12]. Such research, though logistically challenging, has the potential to advance vaccine safety science to reduce the potential for AEFIs with future vaccines [12,13].

International networks for vaccine safety monitoring that include LMICs, such as the Global Vaccine Data Network (GVDN) and, more recently, the International Network for Special Immunization Services (INSIS), have been successfully established, proven feasible, and resulted in valid data generation using harmonized protocols and procedures among the general population [5,13–15]. Such networks also hold promise for exploring vaccine safety within populations that may be excluded from pre-licensure clinical trials (e.g., pregnant and breastfeeding individuals, children, older adults, people with [multi-]comorbidities) [15]. The benefits of these networks include capacity-building using standardized and internationally recognized methods and tools; access to information, expert clinical and epidemiological support across diverse sites; and increased statistical power to explore associations between vaccines and rare adverse events [5,13,15]. Access to international network data has also helped researchers identify differences in AEFI reporting rates between countries, informing the interpretation and generalization of findings, and identifying potential gaps in surveillance [11]. There is limited research exploring the successes and challenges of vaccine safety networks in LMICs and HICs, particularly in regard to biosample collection in AEFI surveillance, and patient perspectives of experiencing AEFIs and participating in vaccine safety research. The studies with evaluative components have tended to be quantitative [11,15,16] and/or retrospective in focus [11,16,17].

Others offer anecdotal lessons learned [18]. Our collaboration among researchers at the Vaccines and Infectious Disease Analytics Research Unit of the University of Witwatersrand (Wits-VIDA) and Wits African Leadership in Vaccinology Expertise (Wits-ALIVE) programme in South Africa, KEMRI-Wellcome Trust Research Program (KWTRP), and Canadian Special Immunization Clinic (SIC) Network via the International Network of Special Immunization Services (INSIS), presented a unique opportunity to investigate facilitators and barriers to vaccine safety surveillance and research across different population groups and settings.

The study objectives were to use an exploratory, multiple-case study approach to provide detailed qualitative findings about the clinical, social, and structural/system factors that contribute to the success of AEFI surveillance and biosampling of cases and controls across LMIC and HIC contexts, and to characterize factors that facilitate and complicate AEFI surveillance and biosampling.

## Materials and methods

### Study design and approach

This study employed a qualitative multiple-case study approach informed by Interpretive Description [19] to examine three different contexts (South Africa, Kenya, and Canada), which are geographically bounded and had unique experiences and perspectives on COVID-19 vaccine safety surveillance. Interpretive description was chosen as it supports the integration of multiple knowledge forms (e.g., clinical, experiential, contextual) to generate practical insights. This design was considered most conducive to gaining insight into overarching similarities and key differences between contexts to inform policy and program recommendations for strengthening vaccine safety evaluation globally.

### Study participants and recruitment

Three groups of participants were recruited: (1) key informants (KIs) with expertise in vaccine safety surveillance or policy in leadership positions in governmental organizations and/or academia; (2) healthcare workers, clinical teams, and laboratory and research staff (HCW/RS) involved in AEFI surveillance, recruitment, biobanking, or clinical management of AEFI cases; and (3) past participants (PP) in vaccine safety and biobanking studies. Participants were enrolled at three sites: Wits-VIDA in Johannesburg, South Africa; KWTRP in Kilifi, Kenya; and the University of Alberta in Edmonton, Canada.

We recruited participants using a combination of purposive, convenience, and snowball sampling. To address potential referral bias inherent in snowball sampling, efforts were made to purposively seek variation in participants’ professional roles, settings, and prior experiences. To capture a broad range of views on AEFI surveillance and research, KIs were identified from each country and via researchers’ networks using purposive sampling based on their involvement in national and/or international immunization policy, surveillance, and vaccine research in participating countries, as well as in other HICs (Australia, United States) and LMICs. Each site recruited KIs from their own country, and additionally, the Canadian site recruited KIs based outside the three participating countries. Purposive and snowball sampling were used for recruiting healthcare workers and research staff, aiming to ensure broad representation of roles and experience. Past participants in vaccine studies who provided consent to contact for future research were contacted for study participation using purposive sampling to ensure representation of different age groups and genders. Recruitment was conducted at the respective sites through a variety of strategies, including email invitations, phone calls, and mailed letters that described the study’s purpose and provided details on participation.

KIs and PPs were recruited to participate in 30 to 60-minute semi-structured interviews. We chose to conduct interviews with KIs to facilitate scheduling with busy health leaders and learn from their unique experiences, and with PPs to ensure comfort and confidentiality in sharing their personal experiences. Healthcare workers and research staff were recruited to participate in 60 to 90-minute focus group discussions, with up to two focus groups conducted per site. Focus group discussions were conducted to explore shared experiences of frontline staff with complementary roles on AEFI surveillance and research teams. Paper-based or electronic informed consent was required prior to participation. Consent was reconfirmed verbally before starting each interview or focus group.

### Data collection

Interviews and focus groups were conducted by trained research team members from each respective site (Canada: ANA; South Africa: NM, S.Shaik, NN, TQ; Kenya: WO, AH, S.Sang, AM). Participants were recruited at the Canadian site from 11 December 2023 to 29 October 2024, at the South African site from 16 February 2024 to 23 May 2024, and at the Kenyan site from 29 April 2025 to 4 July 2025. Semi-structured interview and focus group discussion guides tailored to each participant group were developed based on a comprehensive literature review, expert consultations, and research team discussions (Supplementary Material 1). Interview and focus group discussion topics with KIs and HCW/RS covered experiences with AEFI surveillance during the COVID-19 pandemic, including policy and procedures, infrastructure, data collection and biosampling practices, institutional supports and barriers, and inter-institutional collaboration. PPs were asked about experiences with COVID-19 vaccine decision-making, vaccination itself, and participating in a vaccine research study. At the Canadian site, interview guides for HCW/RS participants were piloted with two healthcare workers with prior involvement in vaccine research, one of whom had personally experienced side effects from a COVID-19 vaccine, to assess the clarity and relevance of the interview questions. Minor modifications to the interview guides were made based on their feedback. All sites reviewed the interview guide for suitability to the local context.

Sample sizes were established in advance to allow for a broad range of experiences and perspectives to be represented across diverse contexts. Interviews were conducted in person or online via cloud-based communication platforms designed for video conferencing and audio recorded. Data collection continued until each site reached its planned sample size for each participant group. In line with narrative and interpretive qualitative methodologies, the emphasis was on capturing rich, diverse accounts rather than pursuing thematic saturation [19]. Demographic information was also collected from participants: gender, country, years of experience and role for KIs and HCW/RS participants, and age and gender for PPs.

### Data analysis

Interview recordings were transcribed and de-identified by two research team members at each site. Transcripts were uploaded into NVivo software in Canada (version 12.6.1.970) and Kenya (version 15.2.0) and to Dedoose (version 9.0.107) in South Africa for analysis. Braun and Clarke’s six-phase thematic analysis was chosen because it provides a transparent and adaptable analytic framework that supports the systematic, iterative construction of themes from qualitative data. Consistent with contemporary understandings of thematic analysis, this approach recognizes that themes are actively developed through sustained analytic engagement rather than passively emerging from the data, while maintaining flexibility to align with the study’s epistemological and methodological commitments [20,21]. The six-step process included: (1) familiarization, (2) initial code generation, (3) theme development, (4) theme review and refinement, (5) theme definition, and (6) final write-up. Two to three trained analysts at each site conducted coding of transcripts (Canada: ANA, S Siraj; South Africa: S Shaik, TQ, NN; Kenya: WO, AH, S Sang). Discrepancies were resolved through discussion with the full research team across all three sites during iterative analysis meetings. Code books were reviewed and discussed regularly among the sites to identify similarities and differences.

The analysis was guided by Interpretive Description methodology [19]. Rigour was ensured through strategies aligned with trustworthiness criteria, including credibility, dependability, transferability, and confirmability [21]. Credibility was enhanced through prolonged engagement with the data, beginning with multiple readings of transcripts to build a thorough understanding before detailed coding. Dependability was achieved via a comprehensive audit trail, comprising coding logs, analytic memos, and documentation of methodological choices. Confirmability was improved by using reflexive practices, like keeping reflective field notes after each interview to record assumptions, potential biases, and immediate impressions. Transferability was ensured by offering detailed, contextual descriptions of the study setting, participants, and findings, enabling readers to assess its applicability to other contexts. The study adhered to the Consolidated Criteria for Reporting Qualitative Research (COREQ) to improve methodological transparency and thoroughness [22].

To support cross-site analysis and interpretation, an in-person research team meeting was held in Johannesburg, South Africa, from May 6-8, 2025, bringing together investigators and field researchers from all participating sites. During this meeting, teams shared preliminary findings, discussed emerging themes, and reflected on context-specific challenges and methodological considerations. This collaborative engagement enabled comparative reflection, deepened cross-site interpretation, and contributed to refining the thematic structure used in subsequent analysis. The meeting also facilitated alignment in analytic approaches while maintaining contextual specificity. As part of the broader analytic process, a stakeholder power analysis was conducted to map the influence and roles of key actors, including policymakers, healthcare workers, community leaders, and families, in shaping AEFI surveillance and reporting practices across sites [23,24].

### Ethics statement

Ethical approval for the study was obtained from the University of Alberta Human Research Ethics Committee (Pro00136220), IWK Health Research Ethics Board (Project #1028279), KEMRI Scientific and Ethics Review Unit (SERU; Protocol no. 5133) and from the University of the Witwatersrand’s Human Research Ethics Committee (HREC - Medical) (clearance certificate no. M230777). Informed consent was required. To preserve confidentiality, participants were assigned a unique ID based on their participant group (KI, HCW/RS, PP), participant number, and study site (Kenya [KE], South Africa [SA], Canada [CA], Canada/International [CA/INT] for international KIs interviewed from Canada).

## Results

### Participant demographics (Table 1)

Across all sites, the team conducted 18 KI interviews, five focus group discussions involving 37 healthcare workers and research staff, and 27 interviews with past research participants. KIs reported between 2.5 and over 30 years of experience in their current roles. Most participants were female. The HCW/RS group included both clinical and laboratory research staff with experience ranging from 1 to over 20 years. Past research participants ranged in age from 22 to 74 years.

**Table 1.**
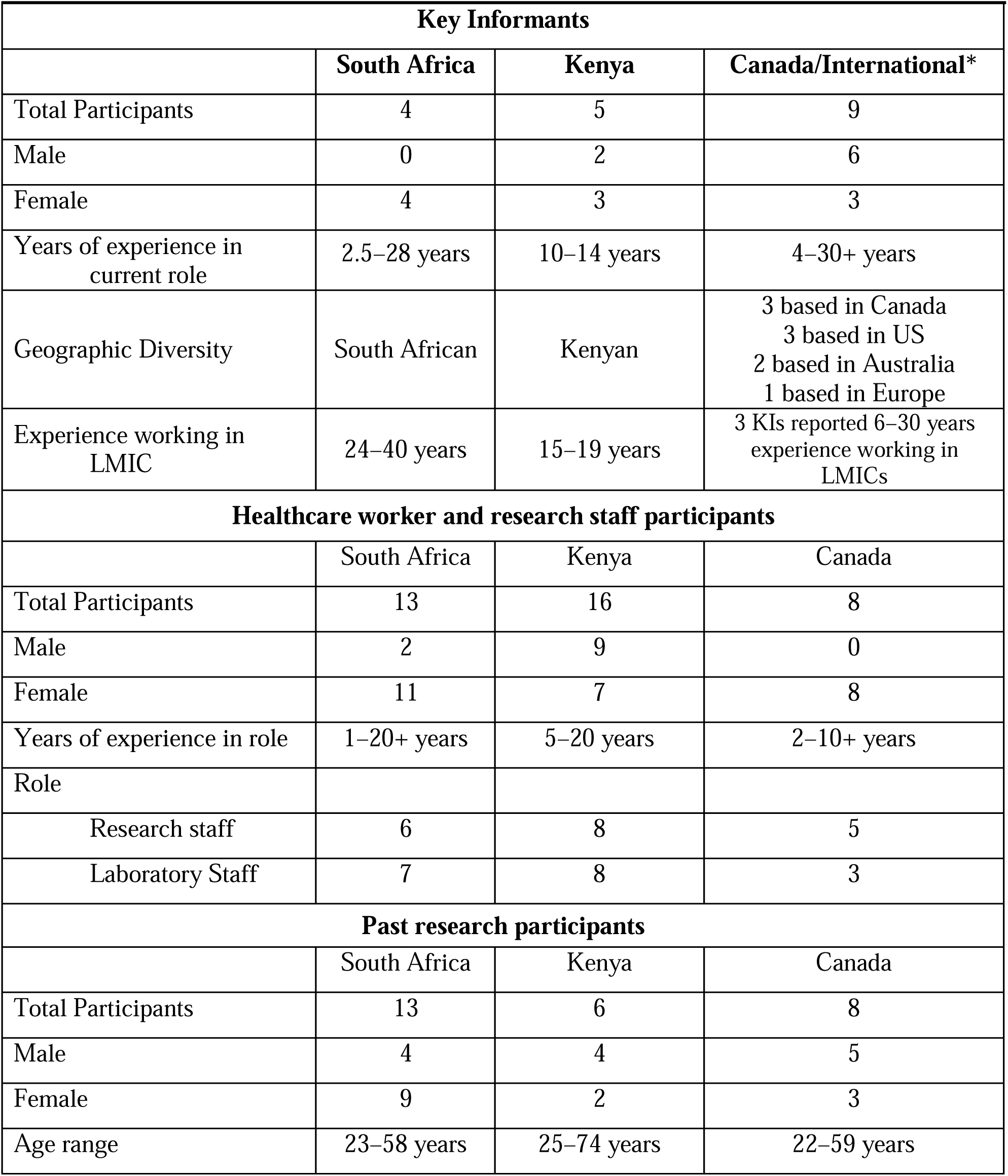
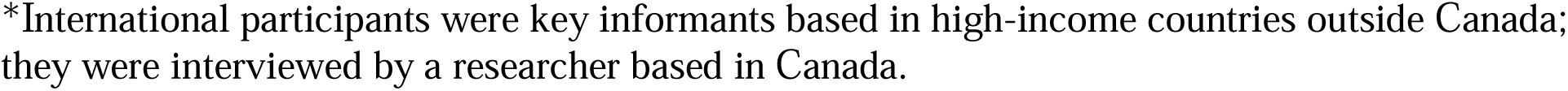
Demographic Information of all research participants across sites.

| **Key Informants** |  |  |  |
| --- | --- | --- | --- |
|  | **South Africa** | **Kenya** | **Canada/International\*** |
| Total Participants | 4 | 5 | 9 |
| Male | 0 | 2 | 6 |
| Female | 4 | 3 | 3 |
| Years of experience in current role | 2.5–28 years | 10–14 years | 4–30+ years |
| Geographic Diversity | South African | Kenyan | 3 based in Canada<br>3 based in US<br>2 based in Australia<br>1 based in Europe |
| Experience working in LMIC | 24–40 years | 15–19 years | 3 KIs reported 6–30 years experience working in LMICs |
| **Healthcare worker and research staff participants** |  |  |  |
|  | South Africa | Kenya | Canada |
| Total Participants | 13 | 16 | 8 |
| Male | 2 | 9 | 0 |
| Female | 11 | 7 | 8 |
| Years of experience in role | 1–20+ years | 5–20 years | 2–10+ years |
| Role |  |  |  |
| Research staff | 6 | 8 | 5 |
| Laboratory Staff | 7 | 8 | 3 |
| **Past research participants** |  |  |  |
|  | South Africa | Kenya | Canada |
| Total Participants | 13 | 6 | 8 |
| Male | 4 | 4 | 5 |
| Female | 9 | 2 | 3 |
| Age range | 23–58 years | 25–74 years | 22–59 years |

\*International participants were key informants based in high-income countries outside Canada; they were interviewed by a researcher based in Canada.

Two overarching themes were developed from the study: (1) AEFI surveillance and investigation across context, and (2) vaccine safety research participation and conduct. Each theme has interrelated sub-themes that capture the nuanced dimensions of participants’ experiences and perspectives (Fig 1).

**Figure 1.**
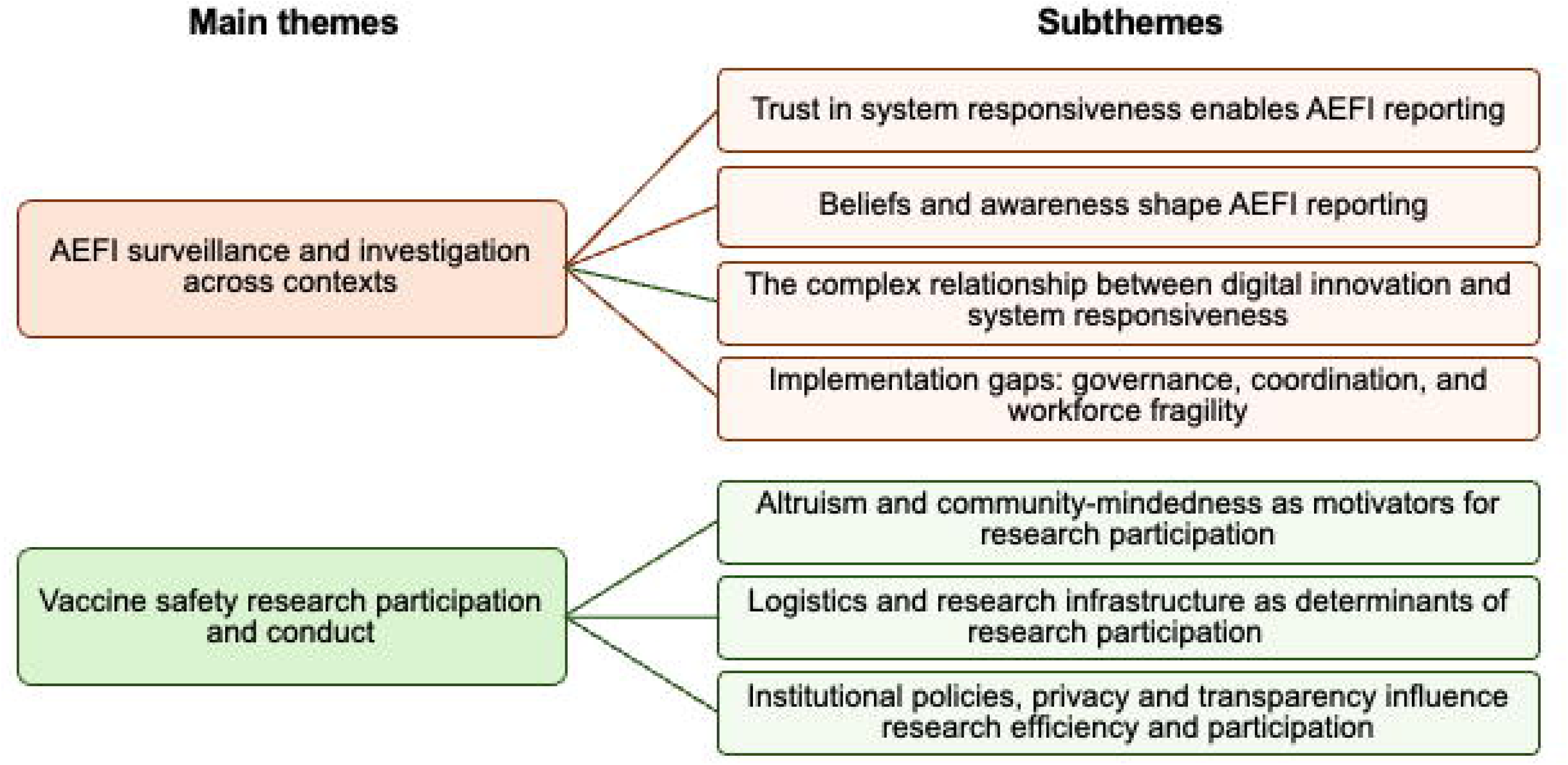
Overview of the major themes and subthemes identified in the study

### AEFI surveillance and investigation across contexts (Table 2)

This theme explores perceptions related to readiness and responsiveness of public health surveillance and healthcare systems in Canada, Kenya, and South Africa to monitor, investigate, and respond to AEFIs. The multi-country findings reveal both shared priorities and site-specific gaps across four subthemes: 1) trust in system responsiveness enables AEFI reporting, 2) beliefs and awareness shape AEFI reporting, 3) the complex relationship between digital innovation and system responsiveness, and 4) implementation gaps: governance, coordination, and workforce fragility.

**Table 2.**
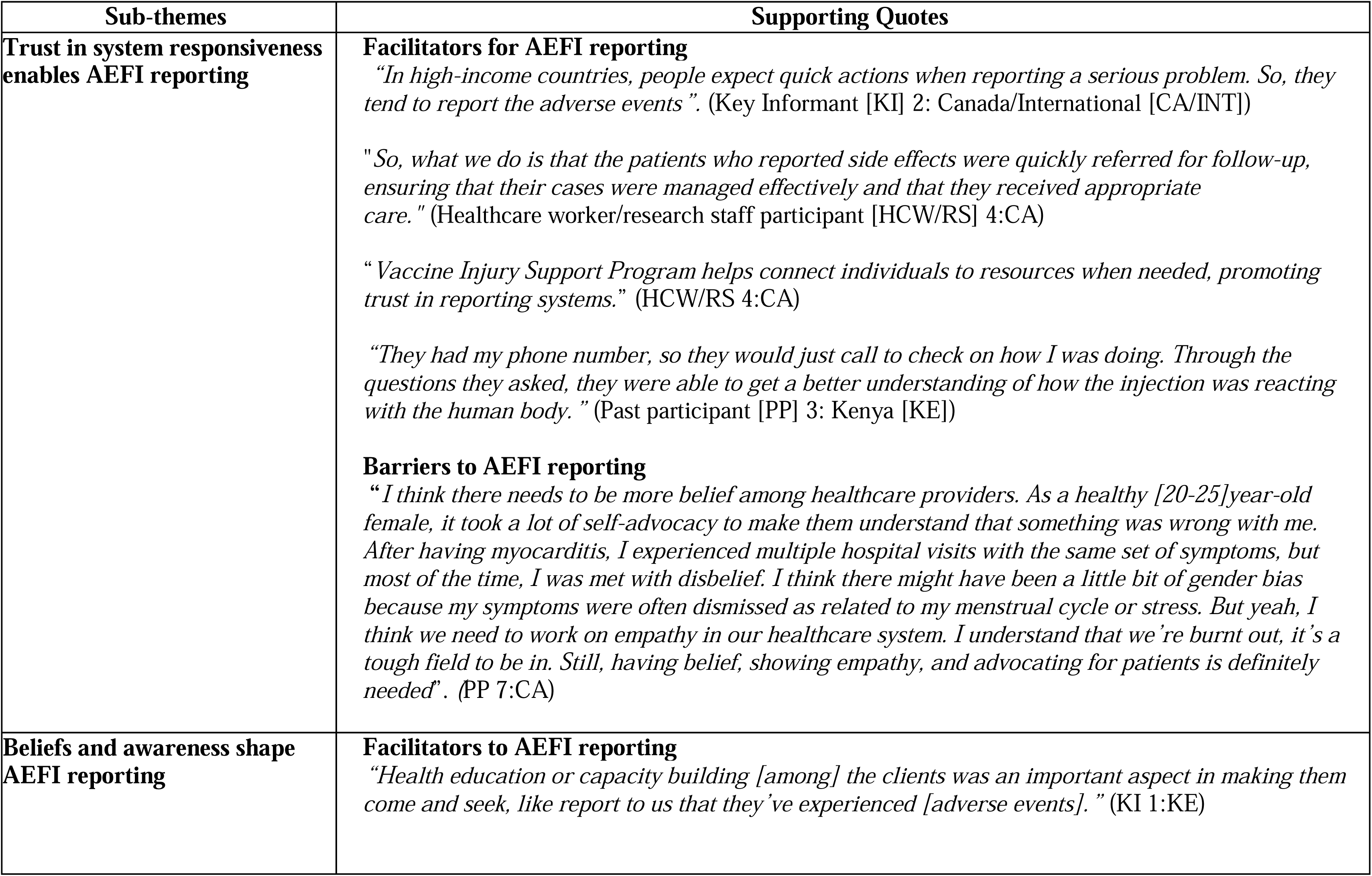
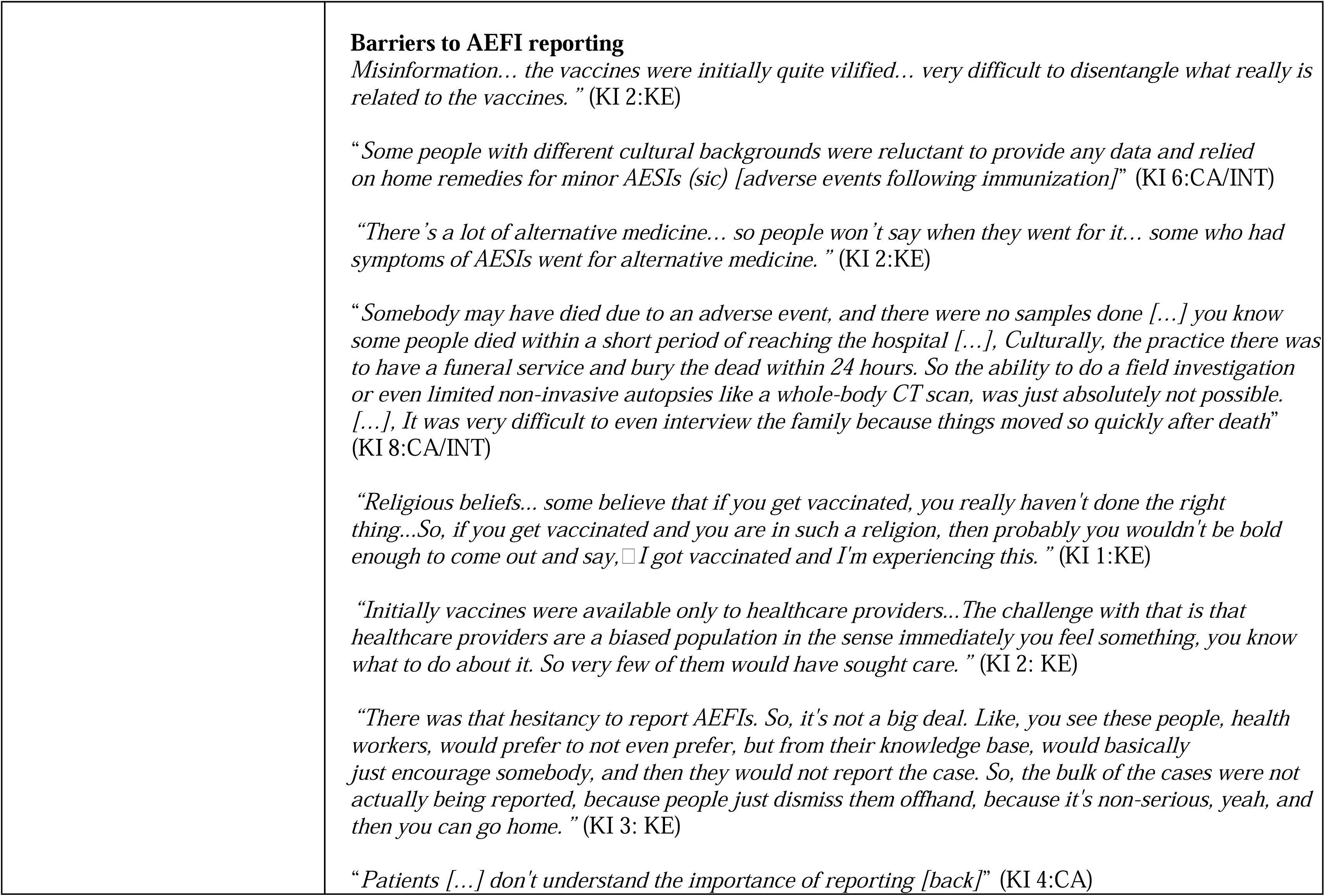
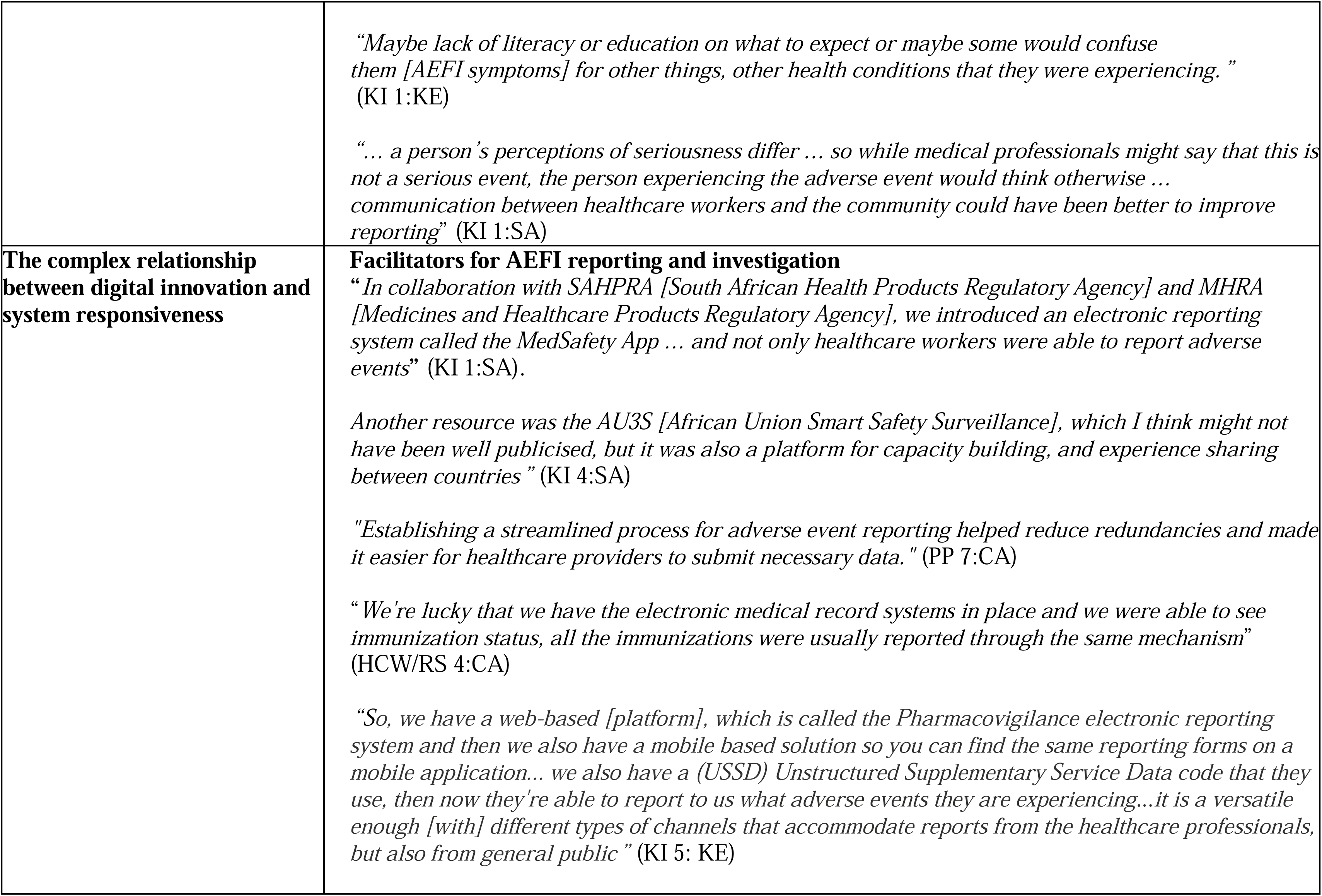
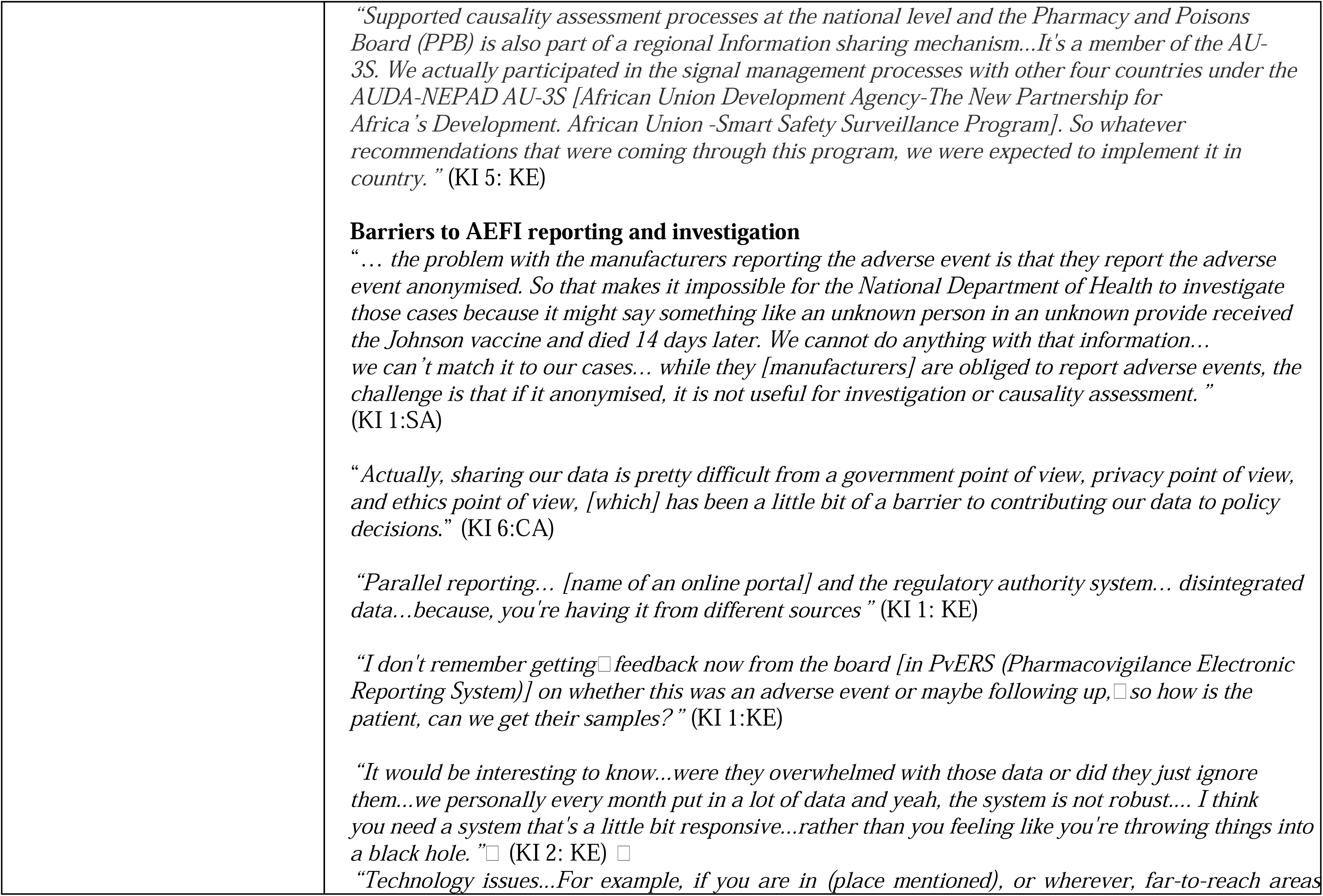
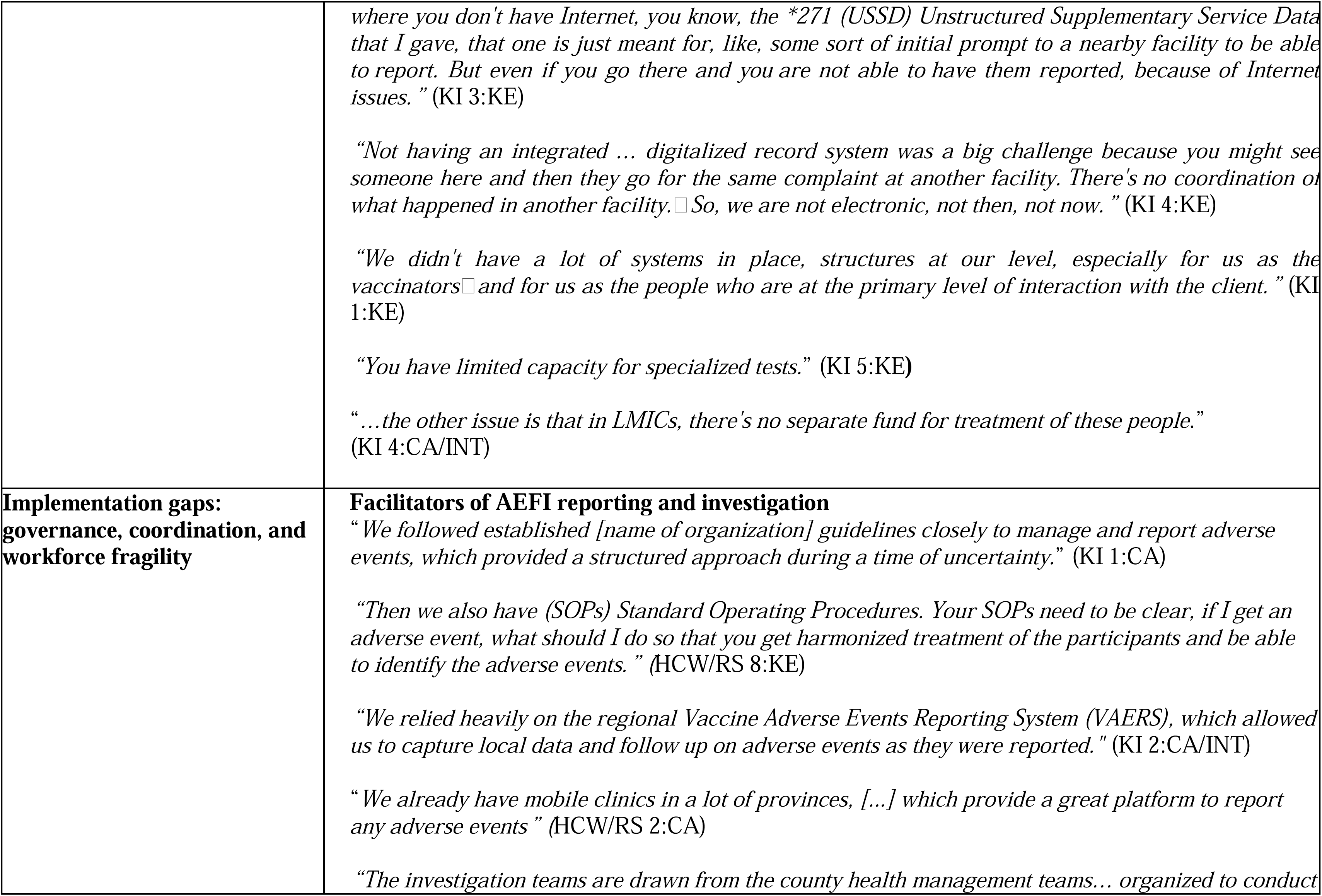
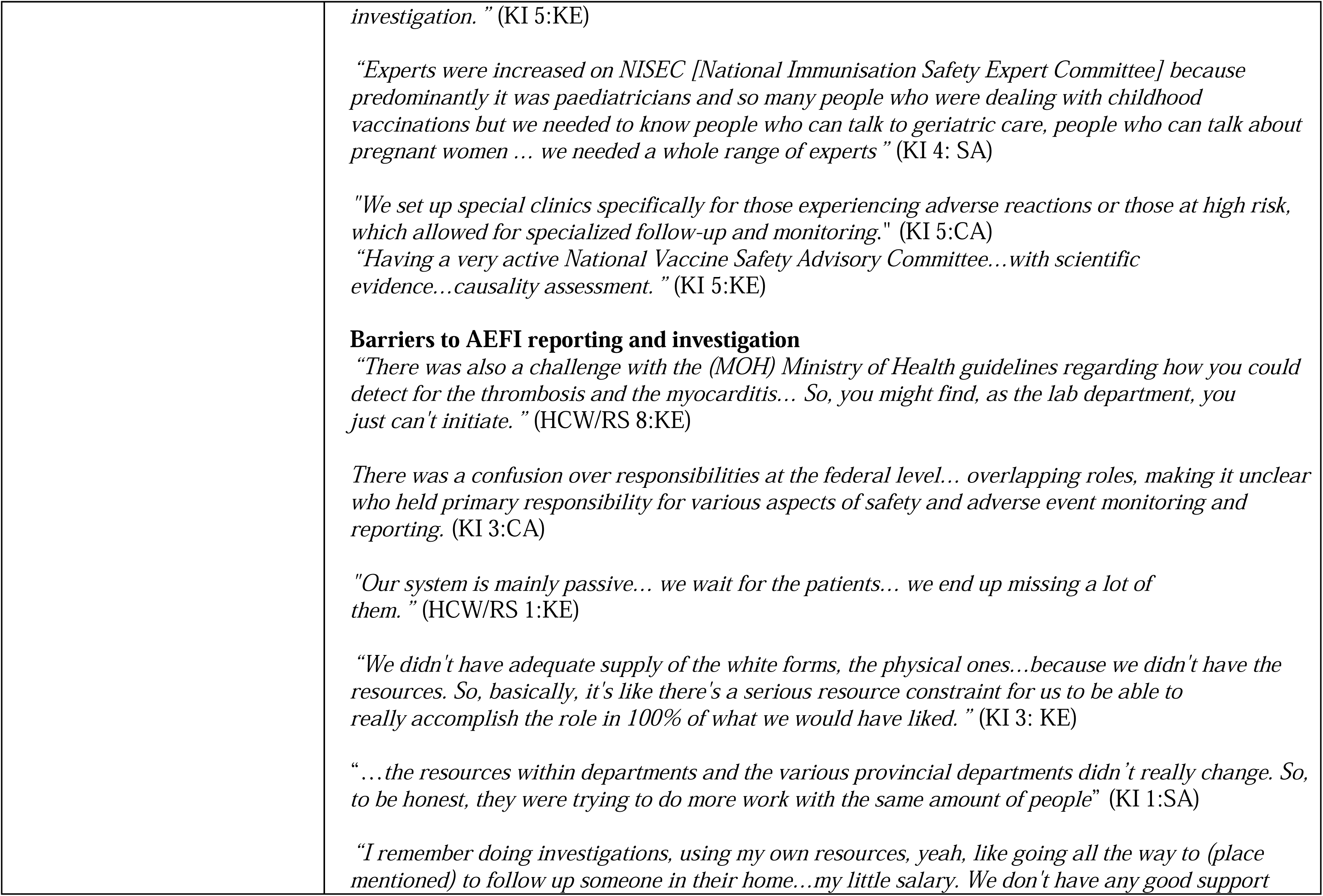
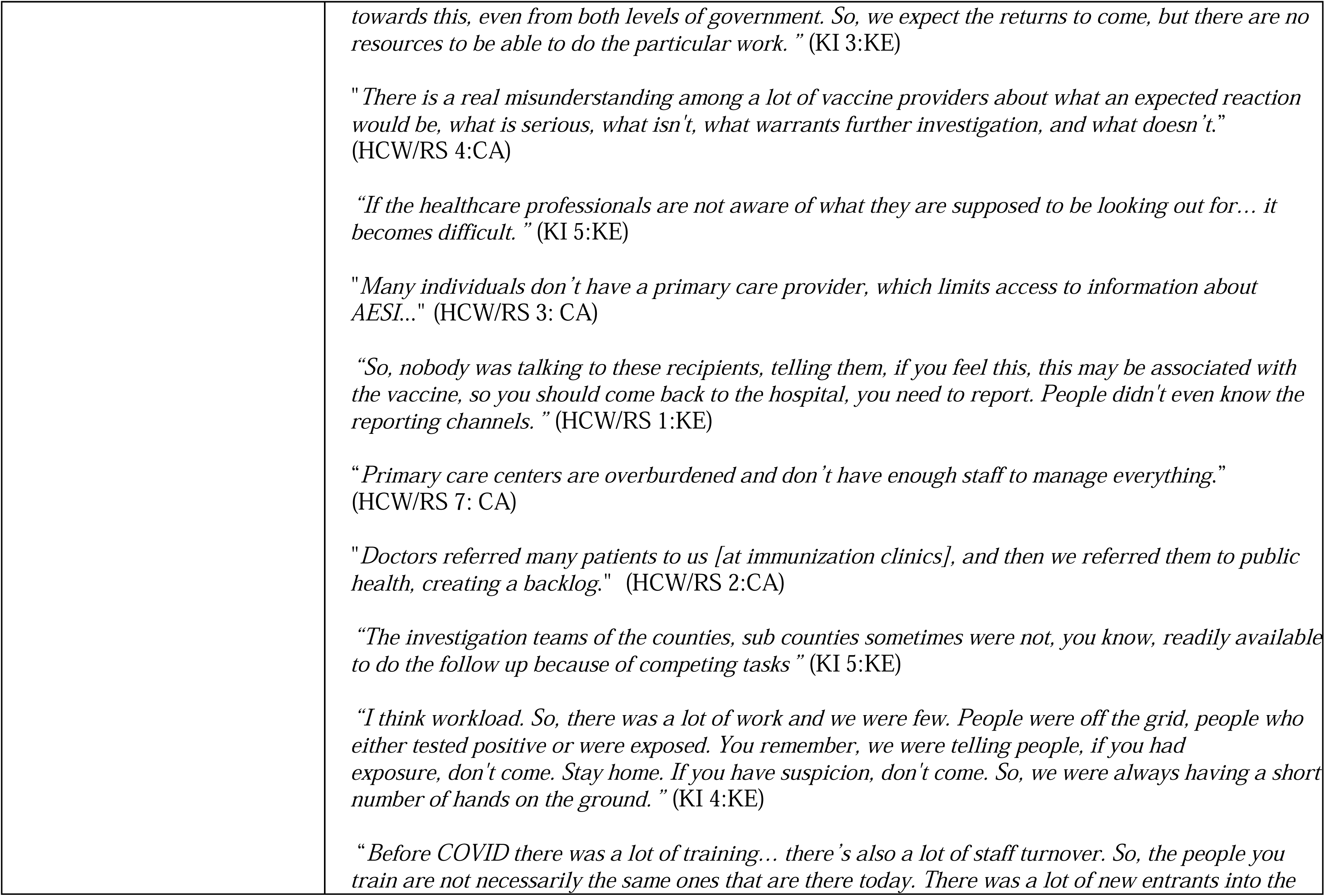
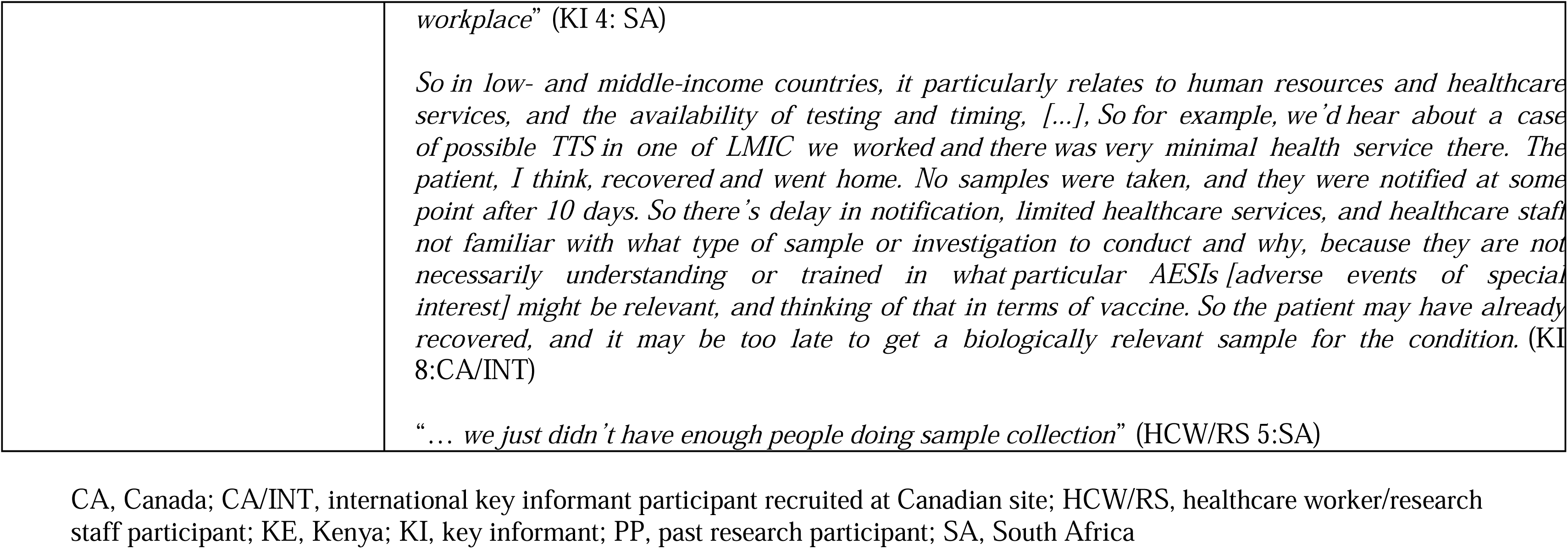
Major theme 1: AEFI surveillance and investigation across contexts subthemes and supporting participant quotes.

#### Trust in system responsiveness enables AEFI reporting

Across contexts, AEFI reporting was described not simply as a technical surveillance task but as a relational process grounded in trust. Participants’ accounts suggest that willingness to report adverse events depends on whether individuals perceive the system as responsive, accountable, and attentive to their experiences. In this way, reporting was framed as a reciprocal exchange: when institutions demonstrate care through follow-up, referral, and communication, individuals are more likely to view reporting as meaningful and worthwhile.

Participants in Canada emphasized that expectations of timely institutional action shape reporting behaviour. As one key informant stated, in settings where people anticipate “*quick actions when reporting a serious problem*,” (KI 2:CA), they are more inclined to report AEFIs. Participants who were healthcare workers described structured referral pathways and follow-up processes that reinforced this expectation, explaining that individuals reporting side effects were “*quickly referred for follow-up, ensuring that their cases were managed effectively and that they received appropriate care”* (HCW/RS 4:CA). Institutional programmes that support individuals after AEFIs were also described as strengthening confidence in the reporting system by signalling that reports lead to tangible support.

A similar pattern was noted in Kenya, where participants highlighted the importance of follow-up contact after reporting an AEFI. Being contacted by healthcare providers was interpreted as a sign of accountability and concern, reinforcing trust in the system. One PP described how follow-up calls helped them feel that their experience was being taken seriously and that their symptoms were being monitored. These interactions contributed to the perception that reporting was not a one-off task, but part of an ongoing relationship between patients and the health system.

At the same time, participants’ narratives also illustrated how trust can be weakened when individuals feel dismissed or are not believed. For instance, a Canadian PP who experienced myocarditis described repeated encounters in which her symptoms were attributed to stress or menstrual factors rather than investigated as a potential vaccine-related adverse event. Her account underscored the importance of empathy and credibility in provider-patient interactions, highlighting how gender bias and clinician dismissal of symptoms can undermine confidence in the system’s responsiveness.

Overall, these findings suggest that trust functions as a key mechanism linking individual experiences to surveillance systems. Follow-up, empathy, and visible institutional action signal that reports are valued and that the system is responsive.

#### Beliefs and awareness shape AEFI reporting

Across sites, KIs shared that AEFI reporting is shaped by how individuals understand, interpret, and frame post-vaccination symptoms. These key informants reflected on patterns they observed in communities and clinical settings, noting that adverse events were often normalized as expected reactions, dismissed as minor, or confused with unrelated health conditions. When symptoms were perceived as routine or not serious, they were less likely to be reported through formal surveillance channels. In this way, reporting was influenced not only by awareness of systems but also by how individuals judged whether their experience was meaningful enough to warrant action.

Beliefs and health practices also shaped decisions about seeking care and reporting. Individuals were sometimes said to rely on “*home remedies*” (KI 6:CA/INT) or turn to “*alternative medicine*” (KI 2:KE) to manage post-vaccination symptoms. This suggests that adverse events might be managed within informal systems of care and thus go undetected even when surveillance mechanisms were available.

Social and religious norms were also described as influencing individuals’ willingness to disclose vaccination-related symptoms. In Kenya, KIs noted that in some religious contexts, accepting vaccination was viewed negatively, making it socially difficult to admit experiencing a vaccine-related health issue. As one KI explained, *“Religious beliefs… probably you wouldn’t be bold enough to come out and say, I got vaccinated and I’m experiencing this”* (KI1: KE). In such contexts, AEFI reporting was not only a health decision but also a socially sensitive act shaped by community expectations and potential stigma. Similar concerns about stigma and reluctance to disclose AEFIs were echoed by KIs recruited fom Canada with experience in LMICs, who observed that social perceptions and personal beliefs could discourage open reporting of symptoms.

Together, these findings show that awareness of reporting systems alone is not enough to ensure that AEFIs are reported. Even when people know that reporting is possible, they may not do so if symptoms are seen as normal, attributed to other illnesses, considered too minor, or deemed socially risky to disclose.

#### The complex relationship between digital innovation and system responsiveness

Across Canada, Kenya, and South Africa, participants described digital technologies as increasingly central to AEFI reporting, suggesting a shift toward more streamlined and accessible surveillance processes. South Africa implemented digital tools such as the MedSafety App, which enabled AEFI reporting by patients and healthcare providers, while data-sharing via the African Union Smart Safety Surveillance (AU-3S) system enabled regional collaboration and comparison of reporting rates between countries. Similarly, in Kenya a digital tool referred to as the Pharmacovigilance Electronic Reporting System (PvERS) facilitated reporting as well as data-sharing via the AU-3S, enhancing regional collaboration. Participants from Canada also reported that streamlined digital reporting systems enhanced AEFI reporting infrastructure.

However, participants’ accounts also revealed that technological solutions did not automatically translate into improved system responsiveness. In Kenya, limited feedback following submissions through PvERS contributed to a perception that reports disappeared into the system, with one participant describing the experience as “*throwing things into a black hole*.” (KI 2:KE). This absence of feedback weakened the perceived value of reporting and undermined engagement with surveillance processes. In both Kenya and South Africa, participants further described limited integration of electronic databases, incomplete vaccine record systems, and restricted access to diagnostic testing, which constrained the ability to validate adverse events and weakened confidence in reporting outcomes.

Data governance and information-sharing challenges further complicated coordination and hampered responsiveness. In South Africa, anonymized manufacturer reports were described as protecting privacy but limiting the ability of national authorities to conduct follow-up investigations or causality assessments. In Canada, stringent privacy legislation and institutional data-sharing restrictions were perceived as slowing collaboration and the timely use of surveillance data. In Kenya, parallel reporting platforms and fragmented data systems were described as producing disjointed information flows, reducing the coherence of surveillance datasets. Across contexts, these findings suggest that data protection and system fragmentation can inadvertently limit the practical usability of surveillance information.

Importantly, participants highlighted that digital tools were embedded within broader structural conditions that shaped their sustainability and effectiveness. While technology can enhance the visibility and flow of AEFI reports, the value of AEFI surveillance depends on ensuring that reports consistently lead to timely investigation, follow-up, and action.

#### Implementation gaps: governance, coordination, and workforce fragility

Across sites, participants acknowledged that policies and procedures for AEFI surveillance were generally well-established and aligned with national and global guidance. However, their accounts highlighted persistent challenges in how these frameworks were implemented, coordinated, and supported in practice. While formal guidelines, expert committees, and reporting pathways were described in Canada, Kenya, and South Africa, participants indicated that gaps in coordination, clarity of roles, and system capacity often limited the ability to translate policy into timely investigation and response.

In Canada, structured institutional guidelines, specialist referral pathways, and support programs were seen as strengthening the formal architecture of surveillance. However, participants described uncertainty regarding the division of responsibilities among federal agencies involved in vaccine safety oversight. While healthcare delivery and case investigation clearly fall under provincial and territorial jurisdiction, ambiguity at the federal level about leadership and coordination of certain surveillance functions was perceived to occasionally slow communication and decision-making. This suggests that even within well-established systems, strong policy frameworks do not always translate into clear operational leadership.

In Kenya and South Africa, national guidelines and expert review mechanisms were also described, including regional investigation teams in Kenya and the expansion of national and provincial immunization safety expert committees in South Africa. Yet participants noted that the absence of clear diagnostic pathways for specific AEFIs, such as myocarditis, and the reliance on passive reporting systems limited case detection and follow-up. Where systems depend primarily on individuals presenting to care, participants felt that adverse events were often missed, indicating that surveillance sensitivity is closely tied to how actively systems seek and verify cases.

Across all settings, workforce limitations emerged as a central constraint linking policy intentions to operational realities. Participants described health systems as overextended, with staff managing multiple responsibilities alongside AEFI reporting and investigation. In Canada, this manifested as heavy care burdens within primary care, where limited time and staffing restricted capacity to support AEFI reporting and follow-up. In LMIC settings, workforce shortages were described as more structural, including insufficient staffing for investigation, limited capacity for sample collection, and high turnover that disrupted continuity and eroded the impact of training. Pandemic-related pressures further intensified these strains, with systems “*trying to do more work with the same amount of people*” (KI 1:SA). Together, these conditions contributed to backlogs, delayed investigations, and reduced system responsiveness.

Overall, participants’ accounts indicate that AEFI surveillance systems are most vulnerable at the level of implementation. Policies, committees, and reporting structures may exist, but gaps in coordination, education, workforce stability, and diagnostic capacity constrain the system’s ability to act on AEFI reports. Surveillance effectiveness, therefore, depends not only on formal governance arrangements but on sustained investment in human resources, training, and clear accountability pathways that enable timely investigation and response.

#### Vaccine safety research participation and conduct (Table 3)

This theme captures participants’ views and experiences related to receipt of COVID-19 vaccination and participation in vaccine safety research. Participants across all three sites recognized vaccine safety research as important for strengthening public confidence and informing immunization programs. However, engagement in research was shaped by a combination of motivational factors, logistical feasibility, institutional research processes, and trust in how data and findings are managed. Three key subthemes were identified: (1) altruism and social influence as motivators, (2) logistics and research infrastructure as determinants of research participation, and (3) institutional policies, privacy and transparency influence research efficiency and participation.

**Table 3.**
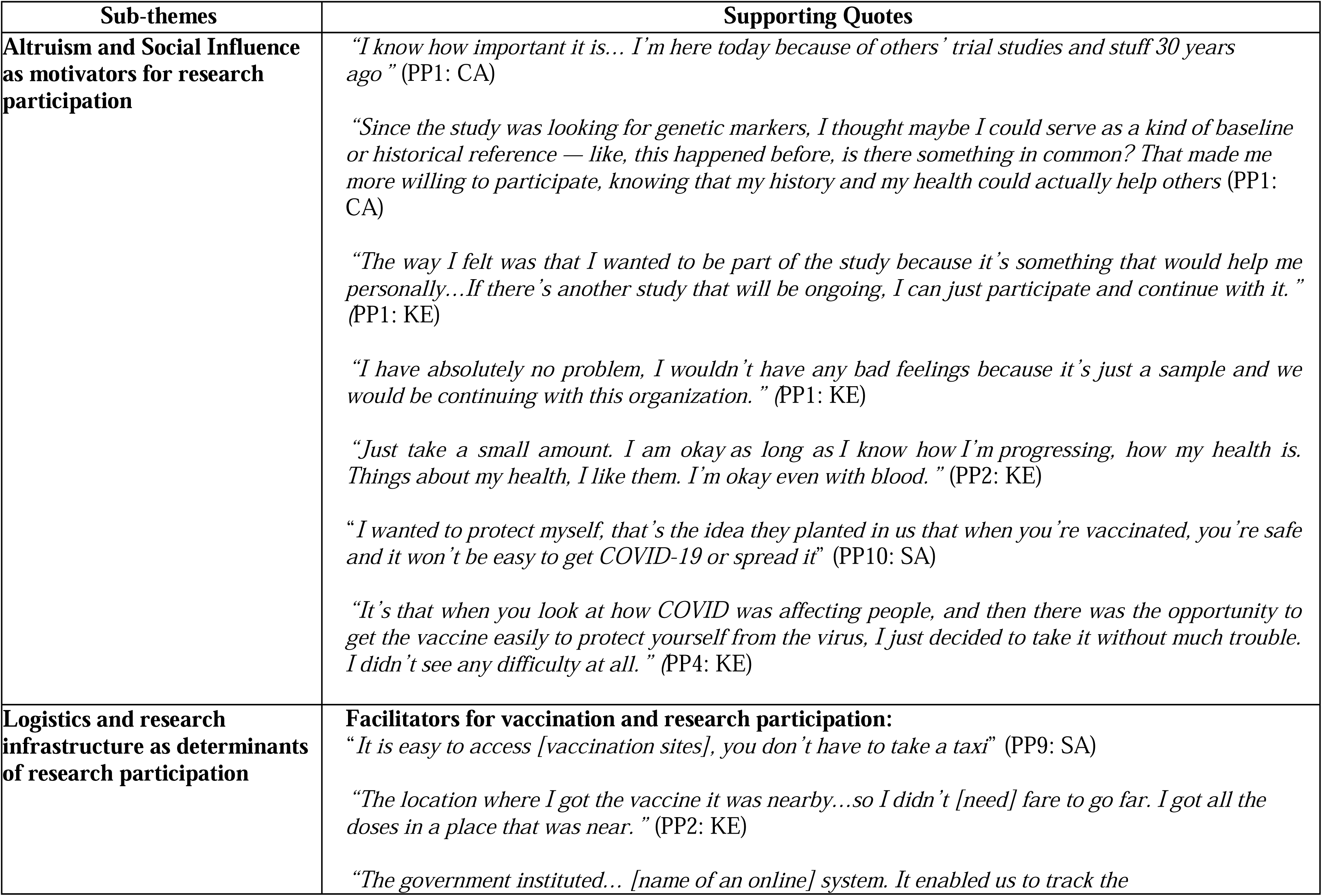
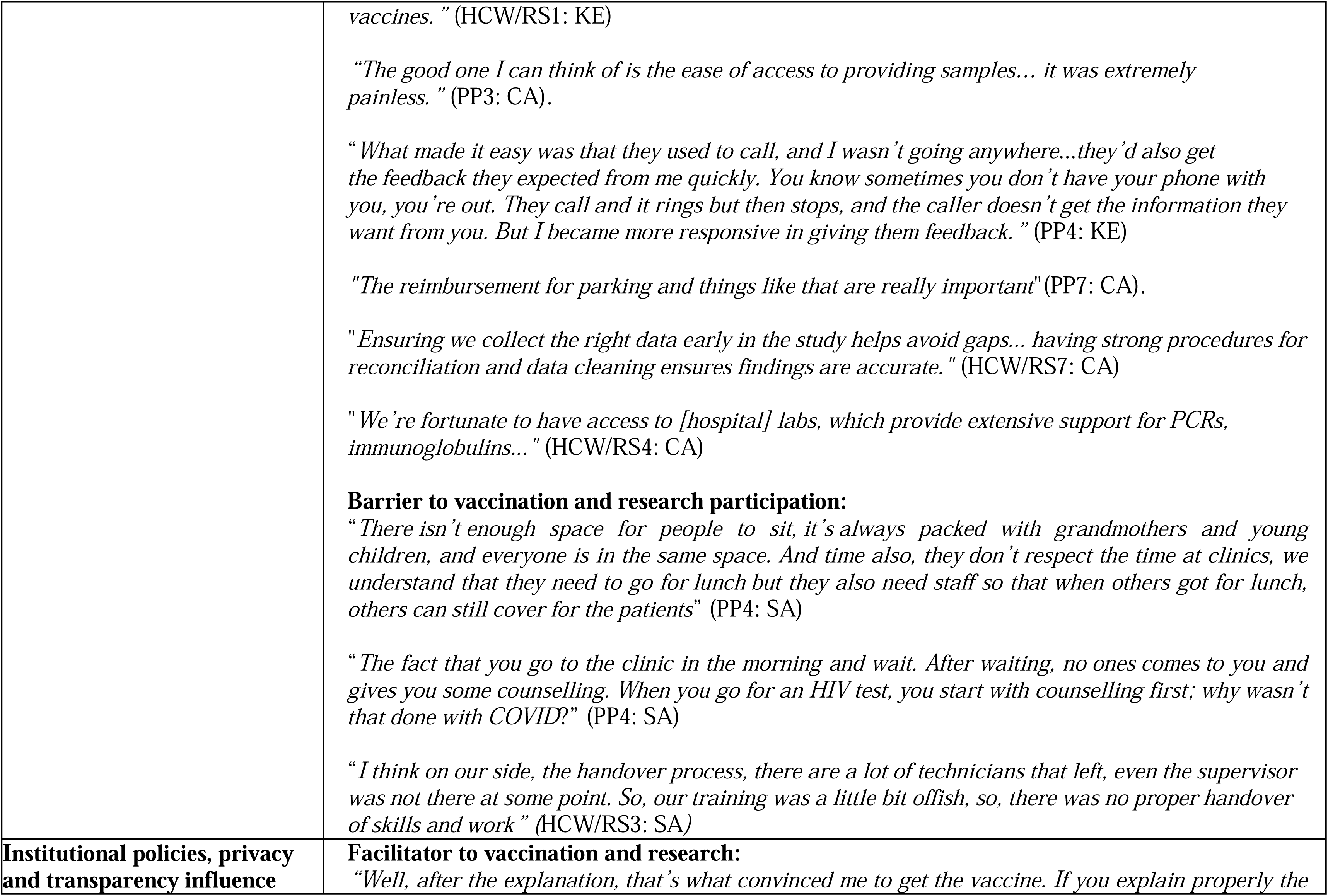
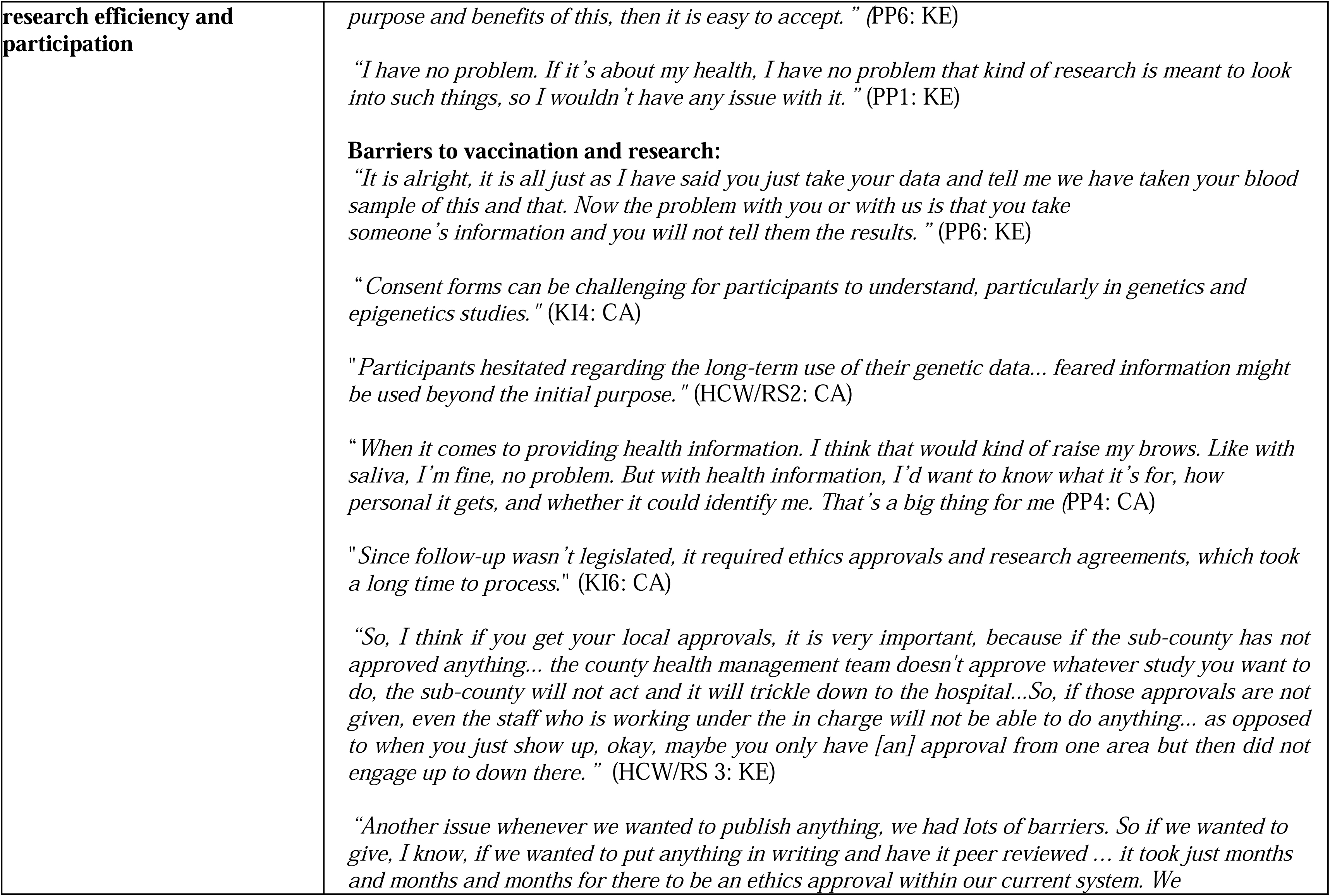
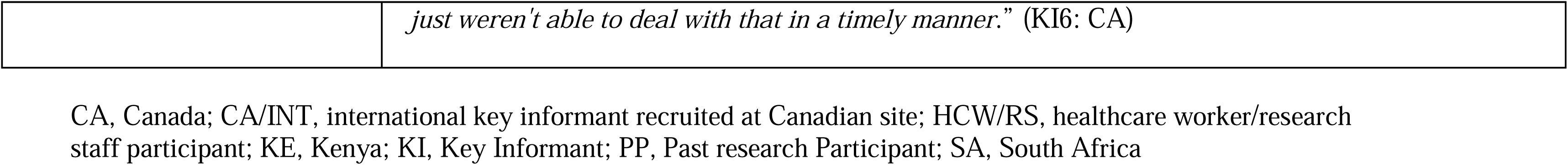
Major Theme 2: Vaccine safety research participation and conduct subthemes and supporting participant quotes.

#### Altruism and community-mindedness as motivators for research participation

Participants from both Canada and Kenya expressed strong motivations to participate in vaccine safety research, driven by a desire to help others and contribute to broader scientific and public health efforts. In Canada, this sense of altruism was often linked to contributing to scientific knowledge, particularly genomic and longitudinal studies that participants believed could benefit future generations. Similarly, Kenyan participants viewed participation positively, describing both personal and communal benefits. Some participants explained that continued involvement in research could provide indirect long-term advantages, such as access to health information, medical follow-up, or ongoing relationships with trusted healthcare and research institutions. These “trusted organizations” included local hospitals, university research programs, and public health agencies that participants perceived as credible, community-oriented, and committed to improving health outcomes. South African participants similarly described gaining knowledge about their illness and receiving clearer information about their health as important motivators for participation.

Perspectives from past participants at all three sites reflected how motivations for research participation were shaped by beliefs about protection, responsibility, and collective well-being. Participants described involvement in vaccine safety studies as a way to contribute to broader community protection and to support public health efforts during a time when COVID-19 was severely affecting their communities. Rather than focusing solely on personal benefit, many expressed a sense of shared responsibility and viewed participation as a meaningful contribution to efforts aimed at improving health outcomes for others. In South Africa, participants also highlighted helping others and advancing medical knowledge as important motivators for joining vaccine safety research. Together, these accounts suggest that motivations for research involvement extended beyond individual gain and were grounded in a broader sense of social responsibility, collective protection, and contribution to public health.

#### Logistics and research infrastructure as determinants of research participation

Logistical factors and research infrastructure played an important role in facilitating research participation in all three contexts. Canadian participants highlighted facilitators such as convenient sample collection processes (providing saliva samples for genomic analysis) and access to hospital laboratories as enabling efficient collection and processing of patient samples. The South African site also noted access to research clinic space and laboratory supported trial conduct. At all three sites, financial compensation and reimbursement for expenses were seen as important enablers for study participants and were appreciated as recognition of the time and effort invested.

Past research participants from South Africa and Kenya also emphasized the importance of physical accessibility of vaccination and research-related services. The proximity and ease of access to vaccination and study sites reduced transportation costs and logistical burden, making both COVID-19 vaccination and research participation more convenient. Conversely, clinic environments that were overcrowded, lacked adequate space, and involved long waiting times without clear communication were described as affecting perceptions of care and the overall acceptability of vaccination services.

Participants further described the role of digital and administrative systems in supporting coordination of vaccination services and research activities. In Kenya, a government-supported online system (Chanjo) was mentioned as enabling tracking of vaccinations and supporting the organisation of vaccine-related services. Standardized data collection and cleaning procedures were noted across settings as important for maintaining research quality. However, logistical barriers were also described. In South Africa, turnover of laboratory staff without adequate handover of responsibilities was reported as disrupting continuity and posing challenges for maintaining consistent research operations.

#### Institutional policies, privacy and transparency influence research efficiency and participation

Concerns related to privacy, consent, and transparency emerged most strongly among Canadian participants, particularly among key informants and healthcare professionals involved in vaccine safety research. They noted that institutional research policies and procedures can affect both the efficiency of research processes and the research participant experience.

In Kenya and South Africa, participants’ experiences with health services also highlighted how institutional practices shaped trust, understanding, and willingness to engage with vaccine and research-related activities. Kenyan participants described positive encounters where clear explanations from healthcare workers influenced their decisions to accept vaccination, noting that when the purpose and benefits of vaccination were explained, it became “*easy to accept*” (PP 6: KE). In contrast, in South Africa, participants compared COVID-19 vaccination experience with other health services (e.g., HIV testing) where individual counselling was routinely provided, suggesting that the absence of individual counselling for COVID-19 vaccination left some feeling insufficiently informed.

Research staff further highlighted the complexity of study consent forms, particularly in specialized areas such as genetic testing, as a barrier to comprehension that could deter participation. Similarly, past participants in vaccine research expressed concerns about sharing sensitive health data when they were unclear about how their information would be stored, used, and protected. Across accounts, transparency, especially regarding future data use and privacy protections, was described as an important consideration influencing decisions to participate in research.

### Integration of stakeholder power analysis

A stakeholder power analysis from this research underscored the importance of harnessing the influence of diverse actors, including politicians, healthcare workers, community leaders, and families to support vaccine communication and AEFI response. Each group holds different forms of decision-making or social capital that can support AEFI response if properly engaged. While institutional actors implement protocols and approve safety decisions, frontline workers build relational trust through direct care. Religious and community leaders, with their emotional and cultural connections, play key roles in information dissemination. Further efforts are needed to engage underserved communities around vaccination, AEFI surveillance and research, using communication channels and tools adapted to the cultures, languages, and education level in those contexts.

## Discussion

This study examined the clinical, social, and structural facilitators and barriers to AEFI surveillance and investigation, as well as vaccine safety research conduct and participation during the COVID-19 pandemic in Canada, Kenya, and South Africa. By engaging key informants, healthcare workers and research staff, as well as past participants in vaccine research across different health system contexts, this study offers critical insights into multisectoral capacities for vaccine safety monitoring and evaluation in high-income and middle-income settings. Our findings highlight several notable facilitators to effective AEFI surveillance and research, as well as several interrelated barriers. Facilitators included robust policies and procedures for AEFI reporting, investigation and follow up, digital tools, and empathic communication, as well as convenient research processes. Barriers included limited trained workforce capacity, weak or fragmented surveillance infrastructure, underfunded systems reliant on external donors, communication gaps in public education, diagnostic constraints for confirming AEFIs, as well as cumbersome research approvals and inadequately addressed privacy concerns.

### AEFI surveillance and investigation across contexts

This study highlights the multifaceted nature of AEFI surveillance and investigation, revealing contrasts in preparedness, system maturity, and contextual needs across settings. In high-income contexts, stakeholder support, regional reporting structures, and consistent communication and training for healthcare providers, as well as specialized clinics for patients experiencing AEFIs, all collectively facilitated AEFI surveillance and investigation. These systems were underpinned by structured digital platforms, electronic medical records, and established pharmacovigilance infrastructure, which in our study, and in prior research, have been identified as enabling timely safety signal detection and response [25]. These findings are consistent with prior studies from HIC and LMIC settings that demonstrated that integrated reporting systems, interoperable data platforms, and strengthened pharmacovigilance networks can improve the timeliness and completeness of AEFI detection, investigation, and regulatory action [15–18,25,26].

Our findings from South Africa are consistent with the work of Sankar and colleagues, who formally evaluated the South African AEFI surveillance system [16]. Their work also highlighted the usefulness of the digital AEFI reporting application (MedSafety App) in facilitating reporting and noted the lack of integration with the existing AEFI surveillance database as a limitation. They identified regional variation in AEFI reporting rates and capacity to review AEFIs, underscoring the need for resources to increase staffing, improve training, and ensure sustained funding. Findings from Kenya identified similar needs for increased funding and resources for AEFI surveillance, as well as the need for clearer guidance and improved access to diagnostic testing to confirm AEFI diagnoses. The AU3S and active COVID-19 vaccine safety surveillance (ACVASS) study, both of which included South Africa and Kenya, offer examples of how coordination of resources and dedicated support can enhance AEFI surveillance and signal detection capacity in LMICs, including for rare adverse events of special interest [15–17]. Both programs enabled integration of AEFI data and strengthening of signal detection capacity, though limited access to digital records, and in the case of ACVASS, a requirement for patient consent to access data, presented challenges to complete case identification [15–17].

Across all settings, there is a clear need for integrated communication strategies and tools including digital applications and portals, online resources, training modules, and targeted media campaigns to support public and provider understanding of AEFIs and AEFI reporting. Effective messaging must not only clarify what constitutes an AEFI but also strike a balance between risks and benefits of vaccination, avoiding stigma while addressing known safety concerns [27–29].

Stakeholders identified key targets for communication efforts, including youth, healthcare workers and vaccinators, as well as community members, and emphasized the importance of leveraging traditional media, social media, and local leaders to build trust and promote evidence-based health information. This resonates with broader recommendations from LMIC and HIC settings to develop localized, multilingual pharmacovigilance campaigns that incorporate trusted community channels and address gaps in health literacy [30–35].

Moreover, managing surveillance in a rapidly evolving global health environment requires adapting to shifts in international support, such as the contraction of budgets for foreign aid, WHO and GAVI, which have direct implications for vaccine procurement and safety monitoring. Strengthening country-level systems and ensuring sustainable investments in pharmacovigilance infrastructure are critical to maintaining public confidence as new vaccines, such as respiratory syncytial virus (RSV) vaccines, are introduced across the life course [27,28].

### Vaccine safety research participation and conduct

Altruism and community motivation emerged as powerful drivers for both vaccine acceptance and research participation in all three settings, although participant concerns and experiences differed across sites. This is consistent with other research showing that individuals often enroll in vaccine trials out of a sense of civic duty and a desire to restore societal normalcy during public health crises [36–38].

Our findings also revealed that fair compensation and reimbursement for study-related burdens are key to participants’ enrollment and retention in research studies. As reported in other studies, addressing these needs promotes equity, respects participant contributions, and supports diverse recruitment [36,37].

Participants in all three contexts highlighted research governance and ethics review processes as key challenges to research study initiation and knowledge dissemination. Specifically, delays in obtaining ethics and institutional approvals were described as hindering timely study initiation and follow-up of participants experiencing AEFIs. These procedural delays were also reported to complicate the dissemination of research findings. Our experience is echoed in other studies from LMIC and HIC settings that have similarly reported that lengthy ethics approval processes delayed study initiation and even prevented sites from participating in vaccine safety research [5,15,37]. There is a critical need to streamline and harmonize research ethics and institutional approval processes to enable research that is timely and responsive to public health needs. Agreements for reciprocal ethics approvals between review boards could lessen the burden and cost on both research teams and ethics review committees. Development and implementation of standard operating procedures aimed at streamlining procedural timelines and ensuring that research adheres to international standards of data governance, transparency, and reproducibility could support faster initiation of high-quality multi-centre and multi-national studies, particularly in research contexts where institutional capacity may be a challenge [36–39].

Interestingly, research consent forms were identified by both research staff and past participants as not meeting participant needs. On the one hand, the forms were reported to be complex and difficult for participants to understand, while on the other, participants expressed concerns regarding how their personal health information would be stored, used, and protected. While participants’ information needs likely vary between individuals and by study type, these findings suggest a need for simpler consent forms that better address participants’ information needs.

These insights underscore the need for streamlined institutional protocols, clearer consent processes, and robust data governance to foster trust and support ethical, participant-centred research practices. The World Health Organization (WHO) Pandemic Agreement provides a roadmap for addressing many of these challenges by calling on member states to promote research collaboration, build research capacity around the world, and support rapid sharing and dissemination of research protocols, materials, and findings [39]. Our findings provide additional insights to support operationalizing the recommendations in the WHO Pandemic Agreement.

### Recommendations (Table 4)

Based on the findings, we developed recommendations for addressing barriers to AEFI surveillance and research that fall into five broad categories: addressing human resource gaps, enhancing surveillance infrastructure, sustained funding, improving communication with the public and underserved groups, and improving diagnostic capacity. Specific strategies include targeted training for frontline healthcare workers, establishment of dedicated AEFI surveillance units, investment in integrated digital AEFI reporting systems including mobile applications that may be more acceptable to younger individuals and improve access in rural areas, sustained national financing, culturally appropriate multilingual awareness campaigns delivered through trusted community channels, and development of standardized investigation protocols and clinical response teams to improve assessment of people with AEFIs. As revealed in the stakeholder power analysis, effective AEFI surveillance depends on engaging actors who hold different forms of influence, from formal decision-making authority to relational and cultural trust, suggesting that system responsiveness requires coordination beyond the health sector alone [15,26].

**Table 4.**
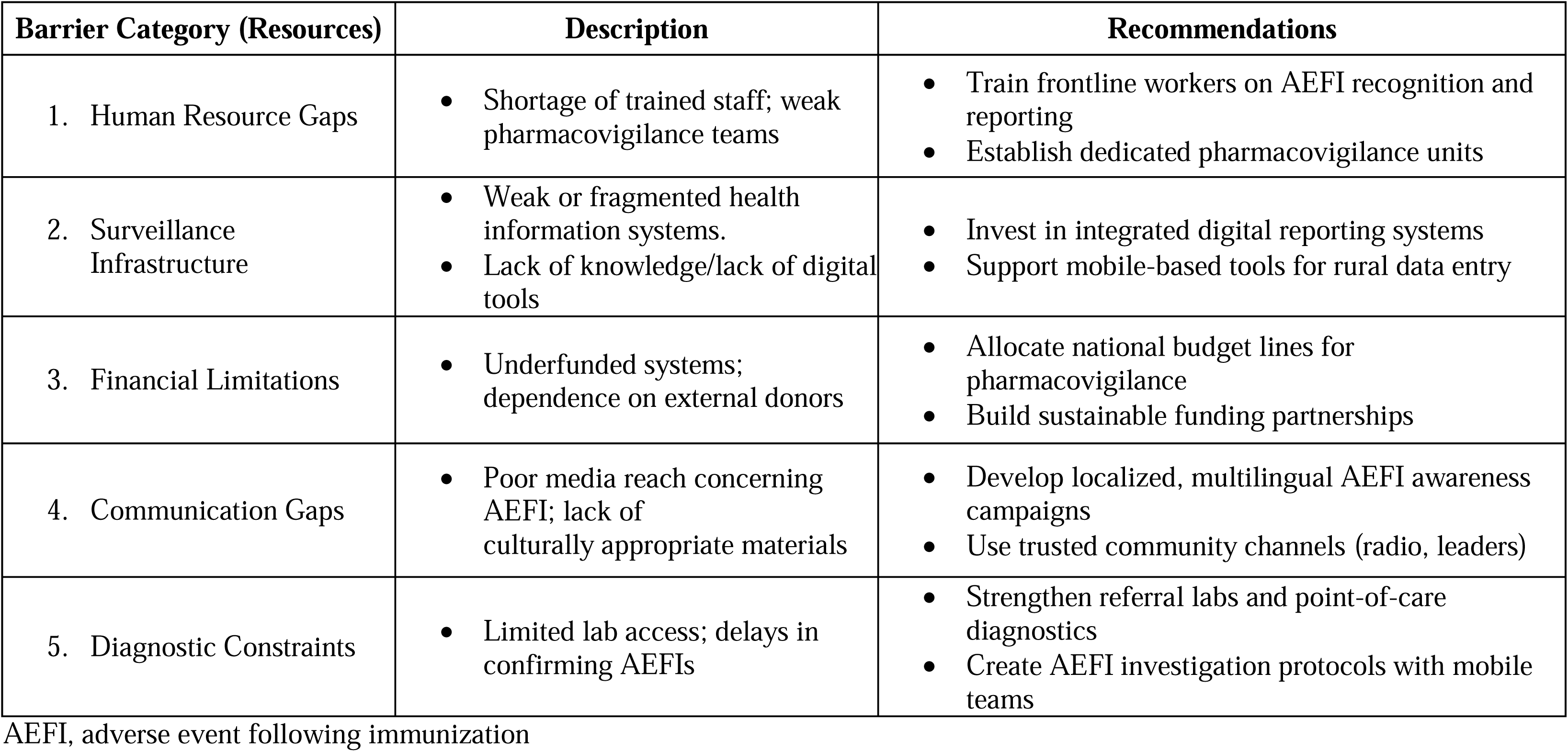
Recommendations for addressing barriers to AEFI surveillance and research.

| **Barrier Category (Resources)** | **Description** | **Recommendations** |
| --- | --- | --- |
| 1. Human Resource Gaps | • Shortage of trained staff; weak pharmacovigilance teams | • Train frontline workers on AEFI recognition and reporting<br>• Establish dedicated pharmacovigilance units |
| 2. Surveillance Infrastructure | • Weak or fragmented health information systems.<br>• Lack of knowledge/lack of digital tools | • Invest in integrated digital reporting systems<br>• Support mobile-based tools for rural data entry |
| 3. Financial Limitations | • Underfunded systems; dependence on external donors | • Allocate national budget lines for pharmacovigilance<br>• Build sustainable funding partnerships |
| 4. Communication Gaps | • Poor media reach concerning AEFI; lack of culturally appropriate materials | • Develop localized, multilingual AEFI awareness campaigns<br>• Use trusted community channels (radio, leaders) |
| 5. Diagnostic Constraints | • Limited lab access; delays in confirming AEFIs | • Strengthen referral labs and point-of-care diagnostics<br>• Create AEFI investigation protocols with mobile teams |

AEFI, adverse event following immunization

### Strengths and limitations

A major strength of this study is its multi-country approach, enabling comparative insights across varied socio-economic and health system settings. Qualitative interviews with key informants, frontline health workers and research staff captured detailed, firsthand perspectives from those directly involved in vaccine safety surveillance, research, and patient care, while incorporating voices of research participants provided additional important insights on improving the patient and participant experience.

Certain limitations are acknowledged. The study used purposive sampling, which may limit the extent to which the results can be applied to regions not represented in each country. Although efforts were made to gather diverse perspectives, certain groups, such as rural populations, youth, or non-English speakers, might have been underrepresented. Moreover, since the study was conducted shortly after the COVID-19 pandemic, the findings may reflect the specific urgency and funding patterns of a global health crisis, which may differ from non-pandemic times.

## Conclusion

This study sheds light on the differing capacities and challenges in AEFI surveillance and vaccine safety research across three distinct health system contexts. While middle-income countries like Kenya and South Africa continue to face critical barriers related to resources, infrastructure, communication, and community engagement, HICs like Canada, despite having more established pharmacovigilance systems, also faced barriers to AEFI surveillance in those areas, which were more pronounced during a public health emergency such as the COVID-19 pandemic. Cross-cutting themes such as the importance of training, digital infrastructure, trust-building, and coordinated stakeholder roles emerged as foundational to effective vaccine safety monitoring and evaluation. As countries adapt to a changing global vaccine landscape, including new vaccine technologies and shifting funding patterns, there is an urgent need to develop locally embedded, ethically grounded, and systemically supported models for AEFI surveillance, investigation and research. These models must prioritize accessibility, equity, and sustainability to build public trust and safeguard vaccine confidence in future immunization efforts.

## Supporting information

COREQ Checklist

## Data Availability

De-identified data are available from the corresponding author upon reasonable written request.

## Acknowledgements

The authors sincerely thank all study participants for sharing their time and experiences. We also acknowledge the expert assistance of Sara Moradipoor and Amanda Bodhi (University of Alberta), and Hannah Munday (Canadian Center for Vaccinology).

†INSIS Investigators who were non-author contributors to this work are:

Robert Chen, Brighton Collaboration, The Task Force for Global Health, Decatur, GA, USA

Nicholas Wood, National Centre for Immunisation Research and Surveillance, Children’s Hospital Westmeade, University of Sydney, Sydney, Australia

Bruce Carleton, University of British Columbia, Vancouver, BC, Canada

Nigel Crawford, Murdoch Children’s Hospital Research Institute, Royal Children’s Hospital, Parkville, VIC, Australia

Ofer Levy, Al Ozonoff, Joann Diray-Arce, Precision Vaccines Program, Boston Children’s Hospital and Harvard Medical School, Boston, MA, USA

C. Buddy Creech, Vanderbilt University Medical Center, Nashville, TN, USA

Paolo Palma, Department of Systems Medicine, University of Rome “Tor Vergata”, Bambino Gesù Children’s Hospital, IRCCS, Rome, Italy,

Steve Black, Global Vaccine Data Network, University of Auckland, Auckland, NZ

## References

1. Nohynek H, Jokinen J, Partinen M, Vaarala O, Kirjavainen T, Sundman J, Himanen SL, Hublin C, Julkunen I, Olsén P, Saarenpää-Heikkilä O, Kilpi T. AS03 adjuvanted AH1N1 vaccine associated with an abrupt increase in the incidence of childhood narcolepsy in Finland. PLoS One. 2012;7(3):e33536. doi: 10.1371/journal.pone.0033536. PMID: 22470453; PMCID: PMC3314666.

2. Greinacher A, Thiele T, Warkentin TE, Weisser J, Kyrle PA, Eichinger S. Thrombotic Thrombocytopenia after ChAdOx1 nCov-19 vaccination. N Engl J Med. 2021;384(22):2092–101. Doi: 10.1056/NEJMoa2104840

3. Schultz NH, Sørvoll IH, Michelsen AE, Munthe LA, Lund-Johansen F, Ahlen MT, et al. Thrombosis and Thrombocytopenia after ChAdOx1 nCoV-19 Vaccination. N Engl J Med. 202;384(22):2124–30.

4. European Medicines Agency. AstraZeneca’s COVID-19 vaccine: EMA finds possible link to very rare cases of unusual blood clots with low blood platelets | European Medicines Agency [Internet]. News. 2021 [cited 2026 Feb 22]. Available from: https://www.ema.europa.eu/en/news/astrazenecas-covid-19-vaccine-ema-finds-possible-link-very-rare-cases-unusual-blood-clots-low-blood

5. Faksova K, Walsh D, Jiang Y, Griffin J, Phillips A, Gentile A, Kwong JC, Macartney K, Naus M, Grange Z, Escolano S, Sepulveda G, Shetty A, Pillsbury A, Sullivan C, Naveed Z, Janjua NZ, Giglio N, Perälä J, Nasreen S, Gidding H, Hovi P, Vo T, Cui F, Deng L, Cullen L, Artama M, Lu H, Clothier HJ, Batty K, Paynter J, Petousis-Harris H, Buttery J, Black S, Hviid A. COVID-19 vaccines and adverse events of special interest: A multinational Global Vaccine Data Network (GVDN) cohort study of 99 million vaccinated individuals. Vaccine. 2024 Apr 2;42(9):2200–2211. doi: 10.1016/j.vaccine.2024.01.100. PMID: 38350768.

6. See I, Lale A, Marquez P, Streiff MB, Wheeler AP, Tepper NK, Woo EJ, Broder KR, Edwards KM, Gallego R, Geller AI, Jackson KA, Sharma S, Talaat KR, Walter EB, Akpan IJ, Ortel TL, Urrutia VC, Walker SC, Yui JC, Shimabukuro TT, Mba-Jonas A, Su JR, Shay DK. Case Series of Thrombosis With Thrombocytopenia Syndrome After COVID-19 Vaccination-United States, December 2020 to August 2021. Ann Intern Med. 2022 Apr;175(4):513–522. doi: 10.7326/M21-4502. Epub 2022 Jan 18. PMID: 35038274; PMCID: PMC8787833.

7. Piché-Renaud P-P, Morris SK, Top KA. A narrative review of vaccine pharmacovigilance during mass vaccination campaigns: Focus on myocarditis and pericarditis after COVID-19 mRNA vaccination. Br J Pharmacol. 2023;89(3):967–981. Doi: 10.1111/bcp.15625

8. Patone M, Mei XW, Handunnetthi L, Dixon S, Zaccardi F, Shankar-Hari M, et al. Risks of myocarditis, pericarditis, and cardiac arrhythmias associated with COVID-19 vaccination or SARS-CoV-2 infection. Nat Med. 2022;28:410–22. Doi: 10.1038/s41591-021-01630-0

9. Klein NP, Lewis N, Goddard K, Fireman B, Zerbo O, Hanson KE, et al. Surveillance for Adverse Events After COVID-19 mRNA Vaccination. JAMA. 2021;326(14):1390–9.

10. Therapeutic Goods Administration (TGA). COVID-19 vaccine safety report - 27-01-2023 [Internet]. COVID-19 Vaccine Safety Reports. 2023 [cited 2026 Feb 22]. Available from: https://www.tga.gov.au/news/covid-19-vaccine-safety-reports/covid-19-vaccine-safety-report-27-01-2023#nuvaxovid-novavax-vaccine

11. van de Munckhof A, Borhani-Haghighi A, Aaron S, Krzywicka K, van Kammen MS, Cordonnier C, Kleinig TJ, Field TS, Poli S, Lemmens R, Scutelnic A, Lindgren E, Duan J, Arslan Y, van Gorp EC, Kremer Hovinga JA, Günther A, Jood K, Tatlisumak T, Putaala J, Heldner MR, Arnold M, de Sousa DA, Wasay M, Arauz A, Conforto AB, Ferro JM, Coutinho JM; Cerebral Venous Sinus Thrombosis with Thrombocytopenia Syndrome Study Group. Cerebral venous sinus thrombosis due to vaccine-induced immune thrombotic thrombocytopenia in middle-income countries. Int J Stroke. 2023 Oct;18(9):1112–1120. doi: 10.1177/17474930231182901. PMID: 37277922; PMCID: PMC10614174.

12. Top KA, Chen RT, Levy O, Ozonoff A, Carleton B, Crawford NW, Creech CB, Kochhar S, Poland GA, Gutu K, Cutland CL. Advancing the Science of Vaccine Safety During the Coronavirus Disease 2019 (COVID-19) Pandemic and Beyond: Launching an International Network of Special Immunization Services. Clin Infect Dis. 2022 Aug 15;75(Suppl 1):S11–S17. doi: 10.1093/cid/ciac407. PMID: 35680552; PMCID: PMC9376276.

13. Diray-Arce J, Chang AC, Moradipoor S, Amodio D, Carleton B, Chang WC, Crawford NW, Karoly M, Hoch A, McEnaney K, Kafil TS, Donthireddy M, Steltz SK, van Haren SD, Angelidou A, Smolen KK, Steen H, Lasky-Su J, Tran H, Liu P, Creech CB, Cutland CL, Petousis-Harris H, Nazy I, Yeung RSM, Kochhar S, Black S, Wood N, Nordenberg D, Palma P, Ovsyannikova IG, Kennedy RB, Poland GA, Ozonoff A, Chen RT, Levy O, Top KA; International Network of Special Immunization Services (INSIS) Members. Longitudinal Meta-cohort study protocol using systems biology to identify vaccine safety biomarkers. Vaccine. 2025 Aug 30;62:127504. doi: 10.1016/j.vaccine.2025.127504. Epub 2025 Jul 26. PMID: 40716144.

14. Carleton B; Global Vaccine Data Network. Genomics of COVID-19 vaccine-induced adverse events [Internet]. [cited 2026 Feb 19]. Available from: https://www.globalvaccinedatanetwork.org/genomics-covid-19-vaccine-induced-adverse-events

15. Cutland CL, Gutu K, Yun JA, Izu A, Mahtab S, Peter J, Ansah NA, Obaro S, Tilahun B, Jambo K, Sow S, Kagucia EW, Chicumbe S, Dlamini T, Browne M, Clothier H, Griffin J, Jiang Y, Lee A, Ghebreab L, Sevene E, et al. Lessons learnt during establishment of COVID-19 active vaccine safety surveillance in nine African countries. Vaccine. 2025 Aug 30;62:127441. doi:10.1016/j.vaccine.2025.127441.

16. Sankar C, Evans S, Meyer JC, Gunter HM, Sekiti V, McCarthy K. Signal Monitoring for Adverse Events Following Immunisation with COVID-19 Vaccines During the SARS-CoV-2 Pandemic: An Evaluation of the South African Surveillance System. Drug Saf. 2025 Aug;48(8):909–922. doi: 10.1007/s40264-025-01547-4. PMID: 40238055; PMCID: PMC12259799.

17. Sankar C, Meyer JC, Schönfeldt M, Gunter H, Dawood H, Sekiti V, Pickard N, Mubaiwa L, Mawela D, Dlamini S, Peter J, Spencer D, Gray C, Patel V, Bamford L, Sehloho T, McCarthy K. Vaccine safety surveillance in South Africa through COVID-19: A journey to systems strengthening. Vaccine. 2025 Feb 6;46:126535. doi: 10.1016/j.vaccine.2024.126535. PMID: 39645433.

18. Fine AM, Goldmann DA, Forbes PW, Harris SK, Mandl KD. Incorporating Vaccine-Preventable Disease Surveillance Into the National Health Information Network: Leveraging Children’ s Hospitals. Pediatrics. 2006;118(4):1431–8.

19. Thorne S. Interpretive description: Qualitative research for applied practice. 2nd ed. Routledge; 2016. doi:10.4324/9781315545196

20. Braun V, Clarke V. Using thematic analysis in psychology. Qual Res Psychol. 2006;3(2):77–101. doi:10.1191/1478088706qp063oa

21. Lincoln YS, Guba EG. Naturalistic inquiry. Beverly Hills (CA): SAGE Publications; 1985.

22. Tong A, Sainsbury P, Craig J. Consolidated criteria for reporting qualitative research (COREQ): a 32-item checklist for interviews and focus groups. Int J Qual Health Care. 2007 Dec;19(6):349–57. doi: 10.1093/intqhc/mzm042. Epub 2007 Sep 14. PMID: 17872937.

23. Balane MA, Palafox B, Palileo-Villanueva LM, McKee M, Balabanova D. Enhancing the use of stakeholder analysis for policy implementation research: towards a novel framing and operationalised measures. BMJ Glob Health. 2020 Nov;5(11):e002661. doi: 10.1136/bmjgh-2020-002661. PMID: 33158851; PMCID: PMC7651378.

24. Bryson JM. What to do when stakeholders matter: Stakeholder identification and analysis techniques. Public Manage Rev. 2004;6(1):21–53. doi:10.1080/14719030410001675722.

25. Phillips A, Carlson S, Danchin M, Beard F, Macartney K. From program suspension to the pandemic: A qualitative examination of Australia’s vaccine pharmacovigilance system over 10 years. Vaccine. 2021;39(40):5968–81. Doi:10.1016/j.vaccine.2021.07.059

26. Top KA, Macartney K, Bettinger JA, Tan B, Blyth CC, Marshall HS, Vaudry W, Halperin SA, McIntyre P; IMPACT and PAEDS investigators. Active surveillance of acute paediatric hospitalisations demonstrates the impact of vaccination programmes and informs vaccine policy in Canada and Australia. Euro Surveill. 2020;25(25):1900562. doi:10.2807/1560-7917.ES.2020.25.25.1900562.

27. World Health Organization, Regional Office for Europe. Behavioural and social drivers of COVID-19 vaccination: tools and practical guidance for achieving high uptake [Internet]. Copenhagen: WHO Regional Office for Europe; 2021 [cited 2025 Dec 23]. Available from: https://www.who.int/europe/publications/i/item/WHO-EURO-2021-2281-42036-57837

28. Kochhar S, Izurieta HS, Chandler RE, Hacker A, Chen RT, Levitan B. Benefit-risk assessment of vaccines. Vaccine. 2023;41(32):4819–25. doi:10.1016/j.vaccine.2023.07.041

29. World Health Organization. The global smart pharmacovigilance strategy [Internet]. Geneva: WHO; 2025 Nov 7 [cited 2025 Dec 23]. Available from: https://www.who.int/publications/i/item/9789240116757

30. Kadio K, Blake-Hepburn D, Song MY, Karbasi A, Noad EE, Abdi S, et al. Facilitators and challenges in collaboration between public health units and faith-based organizations to promote COVID-19 vaccine confidence in Ontario. Int J Equity Health. 2024;23(1):254. doi: 10.1186/s12939-024-02326-w

31. Rabin BA, Cain KL, Watson P, Oswald W, Laurent LC, Meadows AR, et al. Scaling and sustaining COVID-19 vaccination through meaningful community engagement and care coordination for underserved communities: hybrid type 3 effectiveness-implementation sequential multiple assignment randomized trial. Implement Sci. 2023;18(1). doi:10.1186/s13012-023-01283-2

32. Wariri O, Afolabi MO, Mukandavire C, Saidu Y, Balogun OD, Ndiaye S, et al. COVID-19 vaccination implementation in 52 African countries: trajectory and implications for future pandemic preparedness. BMJ Glob Health. 2023;8(12). doi:10.1136/bmjgh-2023-013073

33. Ekezie W, Igein B, Varughese J, Butt A, Ukoha-Kalu BO, Ikhile I, et al. Vaccination communication strategies and uptake in Africa: a systematic review. Vaccines (Basel). 2024 Nov 27;12(12):1333. doi:10.3390/vaccines12121333

34. Driedger SM, Maier R, Metge C, Katz A, Singer A. “I think of it as planting seeds“: challenging patient-provider discussions about COVID-19 vaccination: a qualitative study. BMC Prim Care. 2025;26(1):326. doi:10.1186/s12875-025-03035-1

35. Melnikow J, Padovani A, Zhang J, Miller M, Gosdin M, Loureiro S, et al. Patient concerns and physician strategies for addressing COVID-19 vaccine hesitancy. Vaccine. 2024;42(14):3300–6. doi:10.1016/j.vaccine.2024.04.025

36. Calia C, Reid C, Guerra C, Oshodi A-G, Marley C, Amos A. Ethical challenges in the COVID-19 research context: a toolkit for supporting analysis and resolution. Ethics Behav. 2020;30(1):60–75. doi:10.1080/10508422.2020.1800469

37. Canario Guzmán JA, Orlich J, Mendizábal-Cabrera R, et al. Strengthening research ethics governance and regulatory oversight in Central America and the Dominican Republic in response to the COVID-19 pandemic: a qualitative study. Health Res Policy Syst. 2022;20:138. doi:10.1186/s12961-022-00933-z

38. Dewrance K, Singh S. Recruiters’ lived experiences in COVID-19 vaccine trials in eThekwini, KwaZulu-Natal. Health SA Gesondheid. 2025;30(1). Available from: https://hdl.handle.net/10520/ejc-health_v30_n1_a3123

39. World Health Assembly. WHO Pandemic Agreement. Geneva: World Health Organization; 2025 [cited 2026 Feb 21]. Available from: https://apps.who.int/gb/ebwha/pdf_files/WHA78/A78_R1-en.pdf

