## Supplemental material for "Strengthening Post-Market Vaccine Safety Surveillance Globally: An Interpretive Description Study from Kenya, South Africa, and Canada"

**Supplementary materials**

**Key Informant Interview Guide**

1. Please tell me about your role in adverse event of special interest (AESI) surveillance or similar active surveillance studies [during the COVID-19 vaccination campaign]
2. Please briefly describe the systems or resources available for AESI surveillance and investigation during the COVID-19 vaccination campaign in your region that were important for your work
   a. What factors facilitated AESI case identification and reporting/data collection?
   b. What factors were barriers to AESI case identification and reporting/data collection?

[explore healthcare system factors, patient factors, cultural influences, data management capacity, etc as appropriate depending on role]

c. Were cases followed up? If so, by whom? How many times?
   i. What factors were barriers to follow up data collection?
   ii. What factors facilitated follow up data collection?

3. Were you involved in developing policies or procedures for collecting samples on patients with AESIs and/or controls? If so, were such procedures implemented for the COVID-19 vaccination campaign?
   a. [if yes to both]: What factors facilitated timely sample collection on AESI cases and/or controls? (i.e., at time of AESI diagnosis and/or specific timepoints)
   b. What were barriers to timely sample collection on AESI cases and/or controls?

[explore healthcare system factors, patient factors, cultural influences, laboratory capacity, as appropriate depending on role]

c. Were cases followed up for repeat sample collection?
   i. What factors were barriers to follow up sampling?
   ii. What factors facilitated follow up sampling?

4. From your perspective, how effective were the responses to vaccine safety concerns with COVID-19 vaccines (e.g., myocarditis and thrombosis with thrombocytopenia syndrome)?
   a. What went well?
   b. What aspects could be improved?
5. Based on your experiences with AESI surveillance during the COVID-19 vaccination campaign, what changes would you like to see to improve vaccine safety monitoring for the next mass vaccination campaign?
   a. What improvements would you like to see to the response to the next vaccine safety signal?
   b. What procedures/processes would help to improve timeliness and completeness of AESI investigation including data and sample collection?

**Healthcare worker/research staff participants Focus Group Discussion Guide**

1. Please tell me about your experience with adverse event of special interest (AESI) surveillance or similar active surveillance studies.

*Case Identification (e.g., identifying a case of AESI), confirming vaccination within risk window for AESI (i.e., “true” cases), and data collection/reporting*

2. What were some of the facilitators to case identification, confirmation of vaccination, and data collection/reporting at your site?

- What factors made case identification, confirmation of vaccination, and data collection/reporting easy for you to do?

3. What were some of the barriers to case identification, confirmation of vaccination, and data collection/reporting at your site?

- What factors made case identification, confirmation of vaccination, and data collection/reporting hard for you to do?

*Successful biosample collection and banking (for cases and controls)*

4. Please tell me about the lab facilities available for sample collection, processing, and storage at your site.

- What kind of equipment did you need to implement standard protocols (e.g., provide the INSIS sample collection protocol with equipment list)? Did you have what you needed?

5. Tell me about the staff involved in sample collection, processing, and storage at your site.
6. What was your experience initiating sample collection and biobanking on AESI cases?

If able to initiate:

- How was your experience recruiting?
- What were facilitators to setting up a sample collection process?
- What were barriers to setting up a sample collection process? How did you address those barriers?

If unable to initiate:

- Do you have experiences with sample collection/biobanking for other surveillance studies?
  - What were barriers and facilitators to successful sample collection and processing in those studies?
- What resources and systems would be needed to set up a sample collection process for AESI cases for the next safety concern at your site?

*Consent (adapted to specific contexts)*

7. Can you please tell me about the process for informed consent for participants in AESI surveillance at your site?
   - What factors made obtaining consent for data collection [to retrieve immunization records] easy to do?
   - What factors made obtaining consent for data collection [to retrieve immunization records] hard to do?
8. Can you please tell me about the process for informed consent for specimen collection for vaccine safety studies or similar surveillance activities at your site?
   - What factors made obtaining consent for sample collection easy to do?
   - What factors made obtaining consent for sample collection hard to do?

*Recommendations*

9. What advice/recommendations would you make for other researchers and public health experts looking to provide successful and timely investigation and biosampling of cases with AESIs following vaccination?

**Past Research study participants Interview Guide**

1. To start out we’d like to find out a bit about your views on vaccines, particularly the COVID-19 vaccine and vaccine safety monitoring.
   a. How did you make your decision about **whether or not** to get the COVID-19 vaccine? (e.g. past vaccination experiences, healthcare system, speed of vaccine testing and availability, pressure from healthcare providers, past experiences with public health crises, family members’ perspectives and beliefs, etc.)
      i. Prompts: [if participant got vaccinated]: tell me briefly about your vaccination experience (e.g. registration, appointment, clarity of information provided, where to go afterwards, etc.).
         - what factors made it easier or harder for you to get vaccinated (e.g. environment, provider, location, access, transportation, availability of healthcare provider who spoke your language, past vaccination experiences, etc.)?
         - [if participant did not get vaccinated]: What were some of the factors that led you to this decision? What factors or supports might have led you to get vaccinated?
2. How confident are you (if at all) in the information provided about COVID-19 vaccination?
3. How did this information influence your decision to get vaccinated or not vaccinated, if at all?
4. Please tell me about your experience with participating in a surveillance or vaccine safety study [provider study name as prompt as appropriate].

- How did you feel about being approached for this study?
- Did being approached affect how you felt about vaccines? About your health/reason you were seen in the hospital? How?

5. How did you decide whether or not to participate in the surveillance or vaccine safety study?

- Who, if anyone, did you consult with to make your decision (e.g., family, friends)?

6. (If applicable,) how did you feel about providing a blood sample for the study?
7. (If applicable,) how did you feel about having to follow-up with the team to provide more information or another blood sample?

- What, if anything, made it easy for you to follow-up with the team?

What, if anything, made it hard?

8. How would you feel if you were asked to participate in another, similar study?

- What if it involved collecting your health information?
- What if it involved blood samples?
- What if it involved follow-up visits for data and/or blood collection?

9. What would you like researchers, public health experts, and healthcare providers to know if they want to do more studies like the one you participated in?
